# Cytotoxic T cells are silenced to induce disease tolerance in human malaria

**DOI:** 10.1101/2021.08.19.21262298

**Authors:** Diana Muñoz Sandoval, Florian A. Bach, Alasdair Ivens, Adam C. Harding, Natasha L. Smith, Michalina Mazurczyk, Yrene Themistocleous, Nick J. Edwards, Sarah E. Silk, Jordan R. Barrett, Graeme J.M. Cowan, Giorgio Napolitani, Nicholas J. Savill, Simon J. Draper, Angela M. Minassian, Wiebke Nahrendorf, Philip J. Spence

**Author notes:** these authors contributed equally (Diana Muñoz Sandoval, Florian A. Bach). these authors share senior authorship (Wiebke Nahrendorf, Philip J. Spence).

## Abstract

Immunity to severe malaria is acquired quickly, operates independently of pathogen load and represents a highly effective form of disease tolerance. The mechanism that underpins tolerance remains unknown. We developed a human re-challenge model of falciparum malaria in which healthy adult volunteers were infected three times over a 12 month period to track the development of disease tolerance in real-time. We found that parasitemia triggered a hardwired emergency host response that led to systemic inflammation, pyrexia and hallmark symptoms of clinical malaria across the first three infections of life. In contrast, a single infection was sufficient to reprogramme T cell activation and reduce the number and diversity of effector cells upon re-challenge. Crucially, this did not silence stem-like memory cells but instead prevented the generation of cytotoxic effectors associated with autoinflammatory disease. Tolerised hosts were thus able to prevent collateral tissue damage in the absence of anti-parasite immunity.

## Introduction

The epidemiology of human malaria clearly shows that immunity develops in two distinct phases. First, individuals acquire protection against severe life-threatening disease and in areas of high transmission this occurs very quickly (often before 12 months of age)^1, 2, 3, 4^. Then after many years of exposure protection against clinical malaria is established, which promotes the transition to asymptomatic infection (usually in adolescence)^5^. This temporal dissociation between clinical immunity and immunity to severe malaria suggests that they are underpinned by different mechanisms of host defense. In agreement, clinical immunity often coincides with control of parasitemia (and can therefore be supported by mechanisms of host resistance) whereas immunity to severe malaria is acquired independently of pathogen load and is a form of disease tolerance^1^. One leading hypothesis suggests that broadly neutralising antibodies that recognise variant surface antigens associated with severe malaria (such as group A/DC8 PfEMP1) could prevent severe disease without affecting total pathogen load^2^. In this scenario, immunity to severe malaria would depend upon the rapid production of antibodies that can specifically eliminate pathogenic variants. At present, there is limited *in vivo* evidence that such broad cross-reactivity can be achieved or that neutralising antibodies can be produced within the first year of life to inhibit cytoadherence and reduce sequestration^6, 7^. The mechanism that underpins disease tolerance in human malaria therefore remains unclear.

An alternative explanation is that the host response to infection is quickly modified to minimise the harm caused by malaria parasites. It is well known that metabolic adaptations are induced during the blood cycle to increase host fitness^8, 9, 10^ and control of inflammation might provide an additional path towards disease tolerance^11^. In support of this argument, inflammation decreases with exposure in children (even at high parasite densities)^12^ and pathogenic immune responses can be silenced to minimise tissue stress and toxicity in mice^13^. Importantly, host control of inflammation as a defense strategy would not rule out a role for variant surface antigens in severe disease. After all, inflammation, tissue damage and hypoxia could create the right conditions for endothelium activation and the selection of pathogenic variants^14^. As such, reducing inflammation may minimise the preferential expansion of parasites associated with severe malaria - this would represent a highly effective route to disease tolerance that would not be influenced by parasite strain or genotype.

Controlled human malaria infection (CHMI)^15^ offers a unique opportunity to investigate mechanisms of disease tolerance *in vivo*. Healthy malaria-naive volunteers are inoculated with live (non-attenuated) parasites and their response is tracked throughout infection by repeated sampling (inc. all-important pre-infection samples); volunteers are inoculated with the same clonal malaria parasite (with known variant surface antigen expression) to remove parasite genotype/phenotype as confounding variables; and infections can be terminated at the same parasitemia to maintain a consistent pathogen load. Importantly, this threshold can be safely set at 5,000 - 10,000 parasites ml^-1^ to challenge the immune system with an enormous antigen load (more than 10^7^ parasites per litre of blood at the peak of infection)^14^. What’s more, adults have a far higher incidence of severe disease than children during a first-in-life malaria episode, which means we can study the most at risk group of individuals^16, 17^. We therefore developed a human re-challenge model of falciparum malaria to track the development of disease tolerance in real-time.

## Results

### The risk of severe malaria decreases exponentially with exposure

We first asked how quickly mechanisms of disease tolerance could be established in an area of endemicity. We re-analysed data from a prospective cohort study undertaken in an area of high transmission in Tanzania^1^. Here the authors performed longitudinal sampling with active case detection in more than 800 infants and recorded each independent infection (including pathogen load and disease severity) from birth to 4 years of age. Crucially, there was no reduction in pathogen load across the study period as measured by microscopy (circulating parasitemia) or HRP2 ELISA (total parasite biomass). These data emphasise the absence of host resistance mechanisms in early life. Nonetheless, the authors described a rapid decrease in the incidence of severe malaria consistent with acquired immunity. To ask how quickly the risk of severe malaria decreased we performed maximum likelihood estimation to select the best model fit for these data (Extended Data Figure 1). We asked whether the risk of severe malaria remained constant; decreased linearly or exponentially with exposure; or decreased suddenly after n infections (where n = 1, 2, 3 etc.). The latter stepwise model performed poorly and was removed from further analysis. In contrast, the linear and exponential models provided a good fit and performed better than a constant risk model. To increase our sample size we repeated this analysis and included cases of moderately severe malaria - we collectively describe these episodes of malaria as complicated as 87.7% led to hospitalisation. Once again, the linear and exponential models provided a good fit with the latter performing better by both log likelihood and AIC. Taken together, our results demonstrate that the risk of severe or complicated malaria is highest during the first infection of life and decreases exponentially thereafter (Figure 1A-B). Mechanisms of disease tolerance are therefore beginning to be established after a single malaria episode.

**Figure 1.**
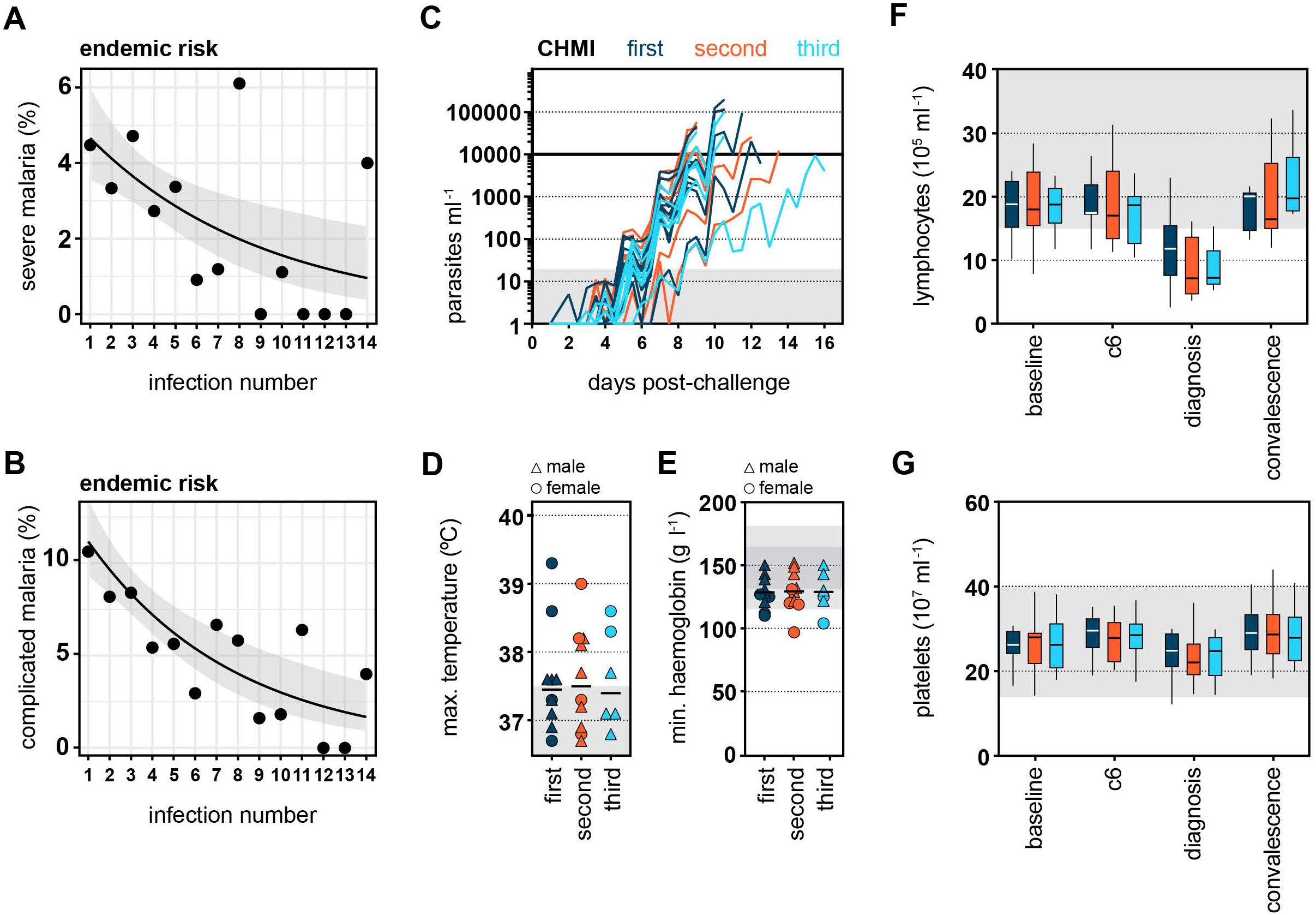
The risk of severe malaria decreases exponentially with exposure. Data were extracted from Gonçalves *et al*.^1^ to examine the frequency of severe (**A**) or complicated (**B**) malaria during the first 14 infections of life in infants living in a hyperendemic setting. We performed maximum likelihood estimation to select the best model fit for these data; the black line shows the best fit and grey shading represents the 95% confidence intervals. In both cases, an exponential decay provided a better fit than either a linear decay or constant risk. In (A) n = 102 severe episodes and in (B) n = 199 complicated episodes of malaria (see Extended Data Figure 1 for case imputation and model performance). (**C**-**G**) Healthy malaria-naive adults were infected up to three times with *P. falciparum* (clone 3D7) by direct blood challenge. Repeated sampling before, during and after each infection allowed us to track the development of disease tolerance in real-time. (C) Parasite growth curves for first, second and third infection; each line represents a volunteer and lines are colour-coded by infection number. Parasite density was measured in peripheral blood by qPCR every 12 hours. The grey box represents the lower limit of quantification (20 parasites ml^-1^) and the treatment threshold of 10,000 parasites ml^-1^ is denoted by the black line. Maximum core body temperature (D) and minimum haemoglobin (E) recorded during each infection (up to 48 hours post-treatment). Each symbol represents one volunteer with a line shown at the median. Grey shading indicates normal range (115 - 165 g litre^-1^ haemoglobin for female and 130 - 180 g litre^-1^ haemoglobin for male volunteers). Lymphocytes (F) and platelets (G) were quantified in circulation the day before infection (baseline), 6 days after challenge (c6), at the peak of infection (diagnosis) and approx. 1 month after drug treatment (convalescence). Boxplots show the median and IQR, and whiskers represent the 95% confidence intervals. Sample size (n) is 10 for first and second infection and n = 6 for third infection. There was no statistically significant difference between groups using a significance threshold of 5% (Kruskal-Wallis test).

### Developing a human re-challenge model of malaria

To identify host adaptations that could quickly reduce the risk of severe disease we developed a homologous re-challenge model of malaria using the most virulent human parasite *Plasmodium falciparum*. Ten healthy malaria-naive adult volunteers were recruited and infected by intravenous injection of parasitised red blood cells during the VAC063A and VAC063B clinical trials; all returned for re-challenge between 4 and 8 months later (during the VAC063B and VAC063C trials, respectively). Six volunteers who took part in both VAC063A and VAC063B were infected for a third time during VAC063C. A blood challenge model was chosen because it standardises the infectious dose, prolongs the period of blood-stage infection (cf mosquito challenge) and removes liver-stage immunity as a possible confounding factor^18, 19^. Importantly, we used a recently mosquito-transmitted parasite line (< 3 blood cycles from liver egress)^20^ since mosquitoes have been shown to reset *Plasmodium* virulence^21^. Volunteers attended clinic the day before infection (baseline), every 12 hours from the day after infection until diagnosis (the peak of infection) and then during the period of drug treatment, which was initiated within 12 hours of diagnosis. These frequent visits allowed for regular blood sampling to construct a detailed longitudinal time-course of each infection. Remarkably, we found that the parasite multiplication rate (and peak parasitemia) were comparable between the first, second and third malaria episode (Figure 1C and Supplementary Table 1). Furthermore, we found no significant change in symptoms or the frequency or severity of pyrexia, anaemia, lymphopenia or thrombocytopenia (Figure 1D-G). These data thus show that healthy adults do not acquire mechanisms of resistance (to reduce their pathogen load) and remain susceptible to clinical malaria. Our homologous re-challenge model therefore recapitulates the key features of endemic malaria in early life.

### Infection triggers a hardwired emergency host response

The absence of anti-parasite immunity means that any change in the host response to infection can not be attributed to a reduced number of circulating parasites. Our model therefore provides the ideal setting in which to investigate mechanisms of disease tolerance. One potential route to tolerance might be to reduce systemic inflammation - this correlates with clinical immunity in endemic regions^12^ but whether it also coincides with immunity to severe disease is not known. To capture the acute phase response we used whole blood RNA-sequencing and DESeq2^22^ to identify differentially expressed genes at diagnosis in first, second and third infection. We found a remarkably similar pattern of interferon-stimulated gene expression regardless of infection number (Extended Data Figure 2A-C). Furthermore, functional gene enrichment analysis showed that the hierarchy of GO terms was near-identical (Figure 2A). These transcriptional signatures were consistent with the rapid recruitment of activated monocytes and neutrophils into peripheral blood, which has been extensively described in naive hosts infected with *P. falciparum*^23, 24^ and *P. vivax*^25, 26^, but it was surprising to see no obvious change upon re-challenge. Nevertheless, by analysing each infection independently it was possible that we were missing important quantitative differences and so we performed direct pairwise comparisons between first, second and third infection. Initially, we compared each pre-infection time-point to identify season-dependent shifts in baseline gene expression - we found zero differentially expressed genes between infections (adj p < 0.05 and absolute fold-change > 1.5). When we then compared each diagnosis time-point to identify adaptations in the host response we again found zero differentially expressed genes (Figure 2B); evidently, the first three infections of life trigger a hardwired emergency response that is not influenced by season or previous exposure.

**Figure 2.**
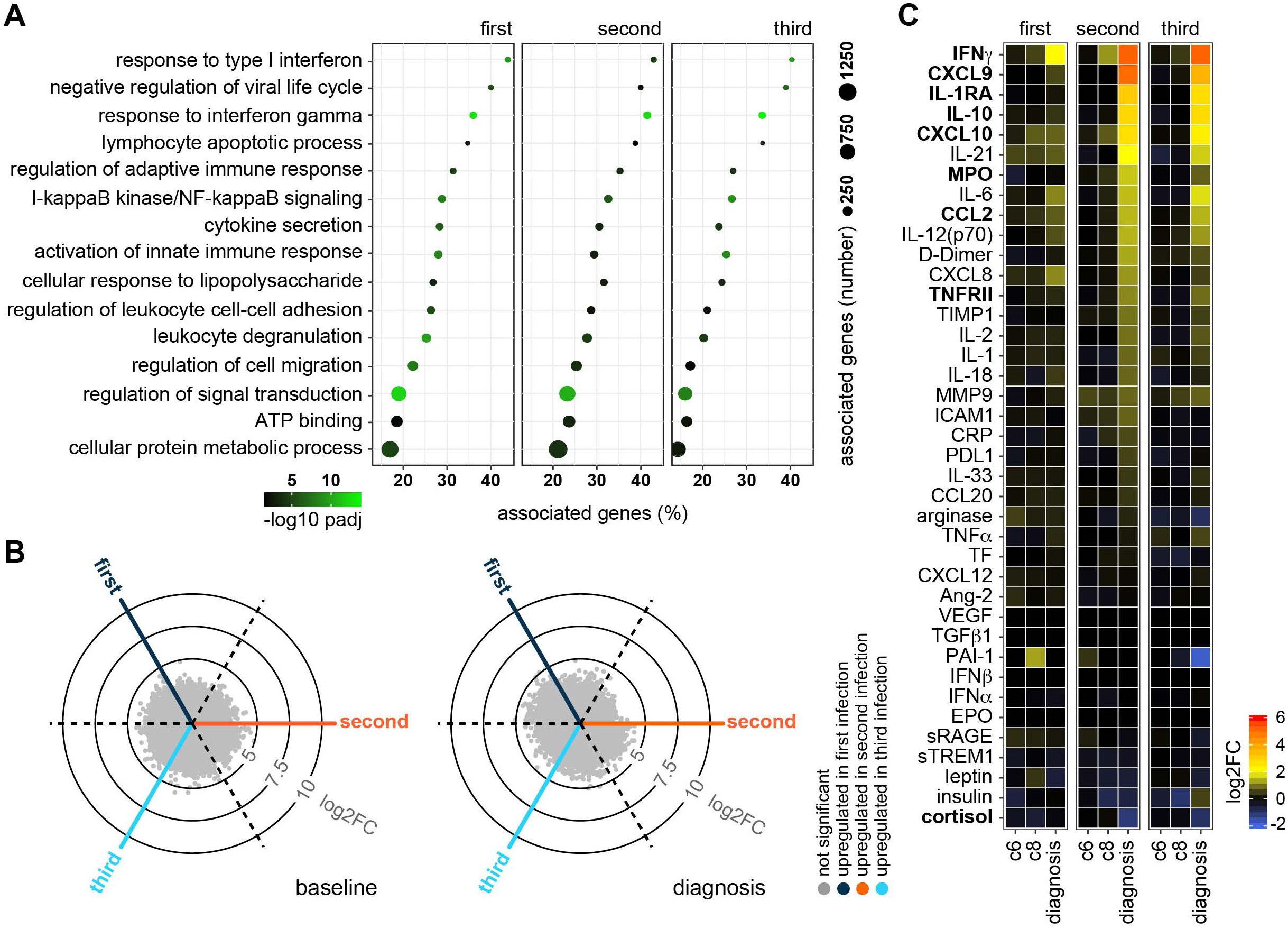
Infection triggers a hardwired emergency host response. (**A**) RNA-sequencing was used to identify differentially expressed genes in whole blood at diagnosis (versus baseline) (adj p < 0.05 and > 1.5 fold-change). ClueGO was then used for functional gene enrichment analysis and placed significant GO terms into functional groups by relatedness. Shown are the leading GO terms from 15 non-redundant groups with the lowest adj p value in first infection. The same GO terms are plotted in second and third infection (note that each infection was analysed independently). (**B**) Radar plots (or 3-way volcano plots) show the number of differentially expressed genes in whole blood between each infection - the left plot compares all baseline samples and the right plot diagnosis. Dashed lines represent the centre-point for each volcano plot and the position of each dot relative to this line shows up- or downregulation. There were no differentially expressed genes in any of the six pairwise comparisons (adj p < 0.05 and > 1.5 fold-change). (**C**) 39 plasma analytes were quantified before and during each infection using a highly multiplexed bead-based assay. The log2 fold-change of each analyte is shown relative to baseline on day 6 and 8 post-challenge (c6 and c8, respectively) and at diagnosis. Analytes are ordered by log2 fold-change and are shown in bold if they varied significantly during both second and third (compared to first) infection (adj p < 0.05 by linear regression with Benjamini-Hochberg correction for multiple testing). In (A-B) n = 10 (first infection), 9 (second infection) and 6 (third infection). v1040 was excluded from RNA-sequencing analysis in second infection because their baseline sample failed QC. In (C) n = 9 (first and second infection) and 5 (third infection). v1040 was excluded from plasma analysis because all samples failed QC.

Nonetheless, host control of inflammation may not be transcriptionally regulated and so we directly measured systemic inflammation at protein level using a highly multiplexed custom bead assay (39 plasma analytes indicative of inflammation, coagulation, oxidative stress and metabolism). By analysing the concentration of each analyte through time we could fit mixed-effects models to test the hypothesis that inflammation was attenuated upon re-challenge (Extended Data Figure 2D). What we actually found, however, was that many of the prototypical products of monocyte and neutrophil activation (such as CXCL10, IL-1RA & MPO) were increased in second and third infection (Figure 2C). The same was true for hallmark cytokines associated with innate lymphoid cell (ILC) or T cell activation (such as IFNγ). Collectively, these data demonstrate that *P. falciparum* triggers a hardwired emergency host response throughout the first three infections of life. And crucially, we find no evidence that systemic inflammation is quickly attenuated.

### A single malaria episode attenuates T cell activation

We therefore moved on to examine adaptive T cells, which are inherently plastic, proliferative and long-lived, and uniquely placed to quickly and permanently alter the host response to infection. The acute phase response to malaria causes extreme lymphopenia leading to a 30 - 70% loss of circulating cells at the peak of infection (Figure 1F). The majority of these are recruited to the inflamed spleen^27, 28^ and so it is difficult to assess T cell activation and differentiation at diagnosis. Instead, we need to analyse T cell activation after drug treatment when the emergency response begins to resolve and T cells return to the circulation^29^. At this time-point, analysing T cell phenotypes in peripheral blood can provide a readout of tissue-specific immune responses. Post-treatment blood samples were not available from the VAC063A or VAC063B clinical trials but were collected during VAC063C. In this trial, we recruited new malaria-naive controls to provide time-matched samples from first infection and infected 11 volunteers contemporaneously (three first infection, two second infection and six third infection) (Supplementary Table 1). As such, we switched to a cross-sectional analysis of VAC063C to incorporate post-treatment time-points.

Six days after drug treatment (designated T6) lymphopenia had completely resolved and all other clinical symptoms of malaria (inc. fever) had receded. Furthermore, markers of systemic inflammation had returned almost entirely to baseline and this was observed in every volunteer regardless of infection number (Extended Data Figure 3A). It was therefore a surprise to find a large transcriptional signature in whole blood at T6 (Extended Data Figure 3B). This signature did not overlap with the emergency response captured at diagnosis and instead the differentially expressed genes had unique functional enrichment terms relating to cell cycle and nuclear division (Extended Data Figure 3C). Remarkably, this proliferative burst was only observed in volunteers undergoing their first infection of life (Extended Data Figure 3D).

Myeloid cells are generally terminally differentiated and do not proliferate after their release from the bone marrow; it therefore seemed likely that our whole blood RNA-sequencing data were capturing the return of activated T cells to the circulation. To explore this further we sorted CD4^+^ T cells with an effector or effector memory phenotype (CCR7^neg^ CD45RA^neg^), which have been shown to dominate the adaptive response to malaria in a naïve host^29^. Sorting was performed on whole blood one day before challenge and six days after drug treatment (Supplementary Data File 1). Analysis of the cell surface markers used for sorting revealed dramatic activation of CD4^+^ T cells in first infection but not second or third infection (Figure 3A) and this striking observation was repeated in a separate CHMI study (VAC069) that infected volunteers with *Plasmodium vivax*, a different species of human malaria parasite that is evolutionarily divergent from *P. falciparum*^30^ (Figure 3B and Extended Data Figure 4A). Our data therefore reveal that the unique feature of a first-in-life malaria episode is fulminant CD4^+^ T cell activation and that this response is attenuated as the risk of severe disease begins to exponentially decrease.

**Figure 3.**
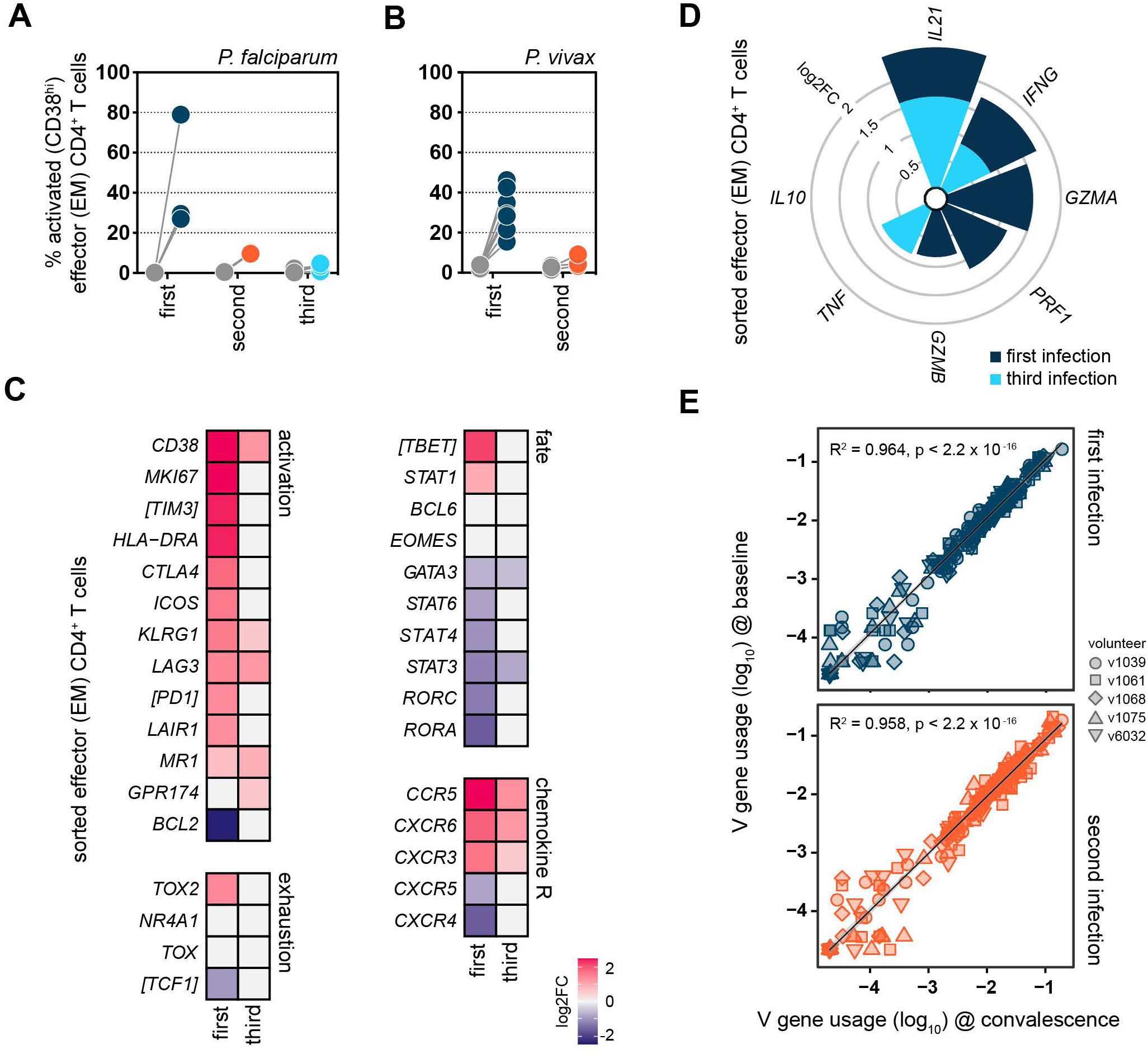
A single malaria episode attenuates T cell activation. (**A**) The percentage of activated CD38^hi^ effector or effector memory (EM) CD4^+^ T cells was analysed by flow cytometry at baseline (grey dots) and 6 days after parasite clearance (coloured dots) during the VAC063C study. Data are shown for volunteers undergoing their first, second or third infection of life (see Supplementary Data File 1 for gating strategies). (**B**) The percentage of activated CD38^hi^ effector or effector memory CD4^+^ T cells at baseline (grey dots) and 6 days after parasite clearance (coloured dots) during the VAC069 study. (**C**-**D**) RNA-sequencing was used to identify differentially expressed genes in flow-sorted effector (effector memory) CD4^+^ T cells (T6 versus baseline) in first and third infection during VAC063C (adj p < 0.05 and > 1.5 fold-change). The heatmaps in (C) show the log2 fold-change of markers of T cell activation and exhaustion and the master transcription factors that shape T cell fate (plus associated chemokine receptors). Non-significant genes are displayed with a log2 fold-change of zero and square brackets indicate that common gene names have been used. The stacked circular bar chart in (D) shows the log2 fold-change of cytokines and cytotoxic effector molecules. (**E**) Correlation between log10 transformed T cell receptor beta variable (TRBV) gene frequency at baseline and 28 days post-challenge (convalescence) showing linear regression (black line) with 99% confidence intervals (grey shaded area). These data represent V gene usage across the entire T cell compartment and were obtained after one or two malaria episodes from volunteers who were subsequently infected for a third time during VAC063C. In (A) n = 3 for first infection, n = 2 for second infection and n = 6 for third infection. In (B) n = 8 for first infection and n = 3 for second infection. In (C-D) n = 2 or 3 for first infection (T6 and baseline, respectively) and n = 6 for third infection (v313 was excluded at T6 because this sample failed QC). In (E) n = 5 in first and second infection (v1065 convalescence samples failed QC).

### T_H_1 polarisation is a unique feature of first infection

To explore the transcriptional landscape of activated CD4^+^ T cells we undertook RNA-sequencing on the sorted effector (effector memory) CD4^+^ T cells obtained from volunteers infected with *P. falciparum*. We found almost 6000 differentially expressed genes at T6 in first infection (adj p < 0.05 and > 1.5 fold-change) and functional gene enrichment analysis showed that these cells were proliferative and had increased their capacity for oxidative phosphorylation (Figure 3C and Extended Data Figure 4B). Furthermore, they had upregulated each of the major costimulatory and inhibitory receptors required to control their fate, and increased their expression of the signature chemokine receptors and transcription factors associated with T_H_1 polarisation. What’s more, the cytokines IFNγ and IL-21 were both strongly induced, which could indicate that infection stimulates double-producers^31^ or that follicular helper T cells were also released from the spleen (Figure 3D).

In third infection, IFNγ and IL-21 were once again significantly upregulated at T6 together with T_H_1-associated chemokine receptors; CD4^+^ T cells are thus transcriptionally responsive to re-challenge (Figure 3C-D). Yet remarkably, the transcription factors that drive T_H_1 differentiation (T-bet and STAT1) were no longer induced, and neither were the costimulatory molecules or inhibitory receptors observed in first infection. We therefore asked what modifies CD4^+^ T cell activation to avert T_H_1 polarisation upon re-challenge. We found no transcriptional evidence that they were quiescent or anergic in third infection (Extended Data Figure 4C) and crucially the transcription factors that epigenetically enforce exhaustion were not upregulated (Figure 3C). Furthermore, there was no evidence for activation-induced cell death (Extended Data Figure 4D) and TCRβ sequencing revealed near-identical repertoires before and after infection, which shows that attenuation was not caused by the clonal deletion of activated T cells after the first or second malaria episode (Figure 3E). There was also no evidence that activated CD4^+^ T cells were diverted towards a regulatory fate; for example, hallmarks of T_R_1 differentiation (such as Eomes and IL-10) were absent (Figure 3C-D).

An alternative explanation could be suppression by conventional regulatory CD4^+^ T cells but our data showed that Tregs were activated in first (not third) infection (Extended Data Figure 4E). It therefore appears that malaria-experienced hosts can launch a specialised adaptive T cell programme that maintains cytokine production without causing extensive activation, proliferation or T_H_1 polarisation. Importantly, we can identify this modified T cell response by transcriptional profiling of whole blood (Extended Data Figure 4F) and it will therefore be possible to identify tolerised hosts in an endemic setting without the need for complex cell isolation protocols.

### Stem-like memory CD4^+^ T cells respond to re-challenge

A major limitation of bulk RNA-sequencing is that there is very little power to detect transcriptional changes if only a small proportion of cells are responding, as seemed to be the case in third infection. The transcriptional landscape of re-activated cells therefore remained largely unclear. To overcome this limitation we used single cell RNA-sequencing to examine individual CD4^+^ T cells, which were sorted at baseline and T6 from three volunteers undergoing first infection and three undergoing third infection; samples were barcoded with oligo-tagged antibodies and superloaded onto the 10X Chromium platform^32^. Three libraries were then prepared and sequenced (cell surface, 5’ gene expression and V(D)J) and after quality control (including doublet exclusion) our dataset contained approximately 25,000 cells per infection with a median of 75,000 reads and 1000 genes per cell (Extended Data Figure 5A-B). Initially, we concatenated all data to identify the heterogeneity of CD4^+^ T cells across the dataset and uncovered 13 unique clusters (7 with a non-naive phenotype) (Figure 4A and Extended Data Figure 5C). We then split the data by volunteer and time-point to examine cluster abundance by linear regression and found that in third infection a single cluster of CD4^+^ T cells (cluster 10) had expanded at T6 (Figure 4B). Importantly, this cluster was transcriptionally high for the canonical activation marker CD38 (Extended Data Figure 5D).

**Figure 4.**
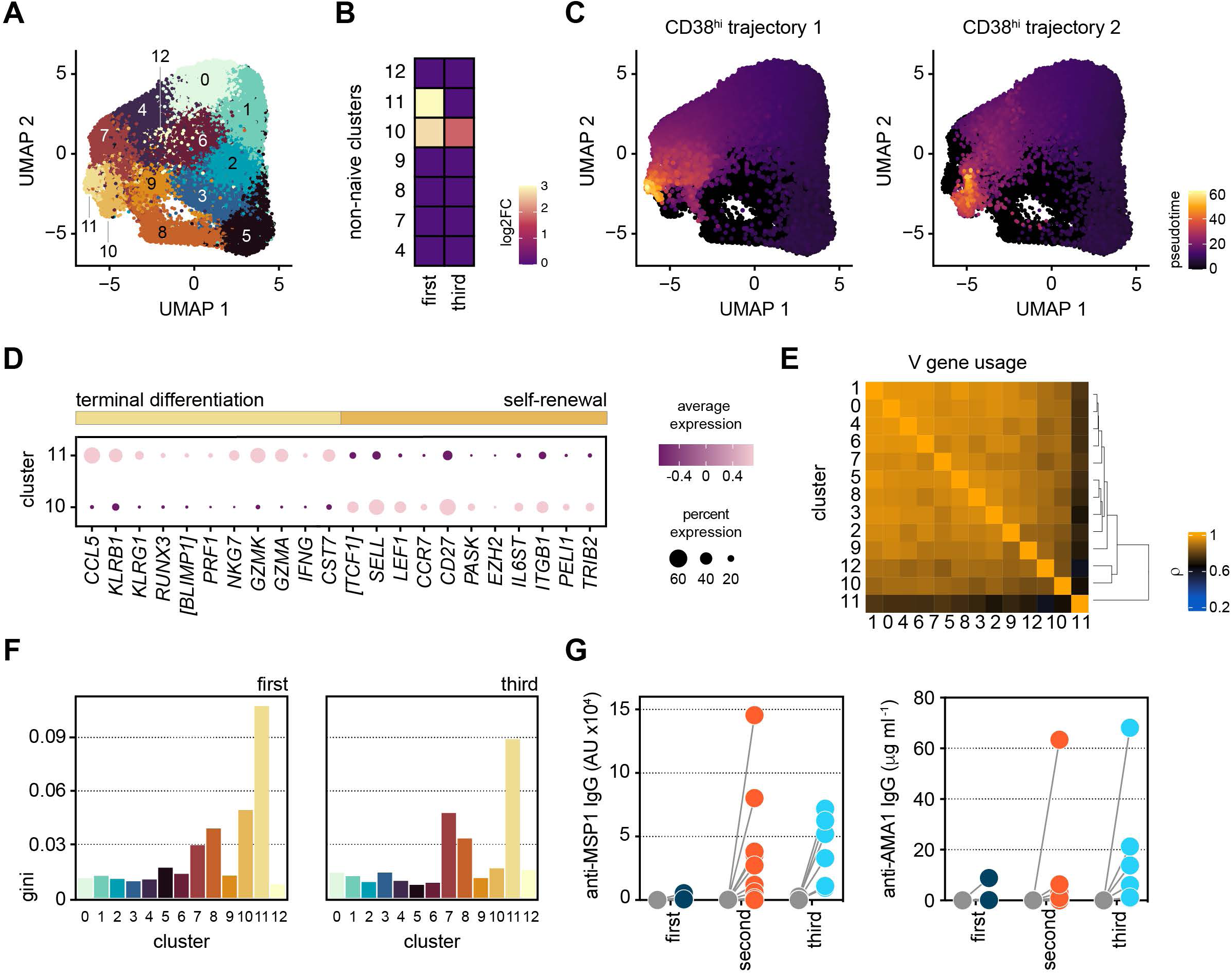
Stem-like memory CD4^+^ T cells respond to re-challenge. Droplet-based single cell RNA-sequencing was carried out during VAC063C on flow-sorted CD4^+^ T cells obtained at baseline and T6 from volunteers undergoing their first or third infection of life. (**A**) Data from all volunteers and time-points was concatenated for UMAP analysis and FlowSOM identified 13 discrete clusters of CD4^+^ T cells across the dataset (each given a unique colour). (**B**) Heatmap showing the differential abundance of non-naive CD4^+^ T cell clusters at T6 (versus baseline) in first and third infection (FDR < 0.05). Note that non-significant clusters are shown with a log2 fold-change of zero and the identification of non-naive clusters is shown in Extended Data Figure 5. (**C**) Dot plot showing differentially expressed signature genes in clusters 10 and 11 (adj p < 0.05). Square brackets indicate that common gene names have been used. (**D**) Trajectory inference with Slingshot revealed clusters 10 and 11 as discrete non-overlapping endpoints of CD4^+^ T cell activation and differentiation (analysis was performed on concatenated data and CD38^hi^ cells were set as the endpoint; Slingshot identified two possible non-overlapping routes). (**E**) Spearman correlation matrix showing shared V gene usage (TRAV and TRBV) across all CD4^+^ T cell clusters (the order of features was determined by unsupervised hierarchical clustering). (**F**) Gini plot showing the equality of V gene usage in each CD4^+^ T cell cluster in first and third infection; zero denotes perfect equality, which indicates a diverse TCR repertoire. (**G**) Class-switched antibodies (IgG) recognising the malaria antigens MSP1 and AMA1 were quantified in serum by ELISA at baseline (grey dots) and one month after challenge (coloured dots). Samples were obtained from volunteers undergoing their first, second or third infection during VAC063A, VAC063B and VAC063C, respectively. In (A-F) n = 3 for first and third infection whereas in (G) n = 10 for first and second infection and n = 6 in third infection.

We therefore examined the signature genes associated with cluster 10 and found enrichment of the transcription factor TCF1 (Supplementary Table 2); this has been associated with circulating T_FH_ cells in a mouse model of malaria^33^ but we found no significant enrichment for PD1, CXCR5 or BCL6 (Extended Data Figure 5E). Instead, we found that cluster 10 was enriched for LEF1, SELL, TRIB2 and PELI1 (Figure 4C), which have all been linked to the maintenance of stem-like properties in T cells^34, 35, 36^. Indeed, we found that BLIMP1, whose induction is required for terminal differentiation, was repressed in cluster 10 (Extended Data Figure 5F) and BLIMP1 is a known target of TCF1^37, 38^. Cluster 10 was also found to expand in first infection but was exceeded by cluster 11, which was similarly high for CD38 but otherwise transcriptionally distinct (Figure 4B-C and Extended Data Figure 5D). Cells in cluster 11 were enriched for signature genes associated with T_H_1 polarisation, terminal differentiation and cytotoxicity, including NKG7 and KLRB1 (CD161), which are hallmarks of rapidly responding short-lived effector cells^39, 40^. Notably, cluster 11 downregulated TCF1 and this could also be seen in our bulk RNA-sequencing data (Figure 3C and Extended Data Figure 5F).

There was no evidence, however, that cells in cluster 11 were descendants of cluster 10 and instead trajectory analysis indicated bifurcation along the TCF1/BLIMP1 axis (Figure 4D and Extended Data Figure 5G). In agreement, TCR V gene usage in cluster 10 shared remarkable overlap with all other T cell clusters but diverged in cluster 11 (Figure 4E). For example, cells in cluster 11 increased TRAV16 / TRBV18 and minimised TRBV5-1 leading to an overall reduction in repertoire diversity (Figure 4F and Supplementary Data File 2). These data are consistent with the rapid clonal expansion of short-lived effectors in first infection, which are silenced upon re-challenge. In contrast, stem-like memory T cells are re-activated and increase their repertoire diversity (to resemble naive T cells) during third infection (Figure 4F). This suggests that cluster 10 is selected for a polyclonal repertoire across multiple malaria episodes and is consistent with reverse TCR evolution^41^. Crucially, the maintenance of stem-like memory appears to support long-lived humoral responses as we observe remarkable stepwise boosting of class-switched IgG antibodies against MSP1 and AMA1 (Figure 4G).

### Cytotoxic T cells are silenced for a minimum 8 months

The terminal differentiation of cytotoxic CD4^+^ T cells has been described in autoinflammatory disease, the tumour microenvironment and chronic (or latent) viral infection^42, 43, 44^. We were nevertheless surprised that our bulk and single cell RNA-sequencing data both suggested that T_H_1 polarised CD4^+^ T cells acquired transcriptional features of cytotoxicity during a first-in-life malaria episode (Figure 3D and 4C). Furthermore, the scale of activation (up to 80% of all circulating cells with an effector or effector memory phenotype (Figure 3A-B)) was staggering, and far exceeded observations made in other human challenge models of acute infection, such as influenza^45^ or typhoidal salmonella^46^. Our data therefore suggested that rather than the response to re-challenge being unusual, the host response to a first infection was excessive and disproportionate. To explore this idea we used mass cytometry so that we could examine more cells, extend our analysis to all major T cell subsets and quantify the heterogeneity of activated cells at protein level. To this end we designed an antibody panel that prioritised key markers of T cell function and fate (Supplementary Table 3).

Whole blood samples were preserved in Cytodelics Stabilisation buffer within 30 minutes of blood draw at baseline, diagnosis and T6 (as well as during convalescence) during VAC063C. As before, we concatenated all data to identify the heterogeneity of T cells across the entire dataset and uncovered 49 unique clusters (Supplementary Data File 3). As expected most of the diversity was observed within the non-naive CD4^+^ and CD8^+^ T cell subsets (Figure 5A). Tracking the frequency of each cluster through time then resolved dynamic changes in the T cell compartment (Supplementary Data File 4) and we performed linear regression on cell count data using edgeR^47^ to identify the differentially abundant clusters at each time-point (FDR < 0.05 and absolute fold-change > 2).

**Figure 5.**
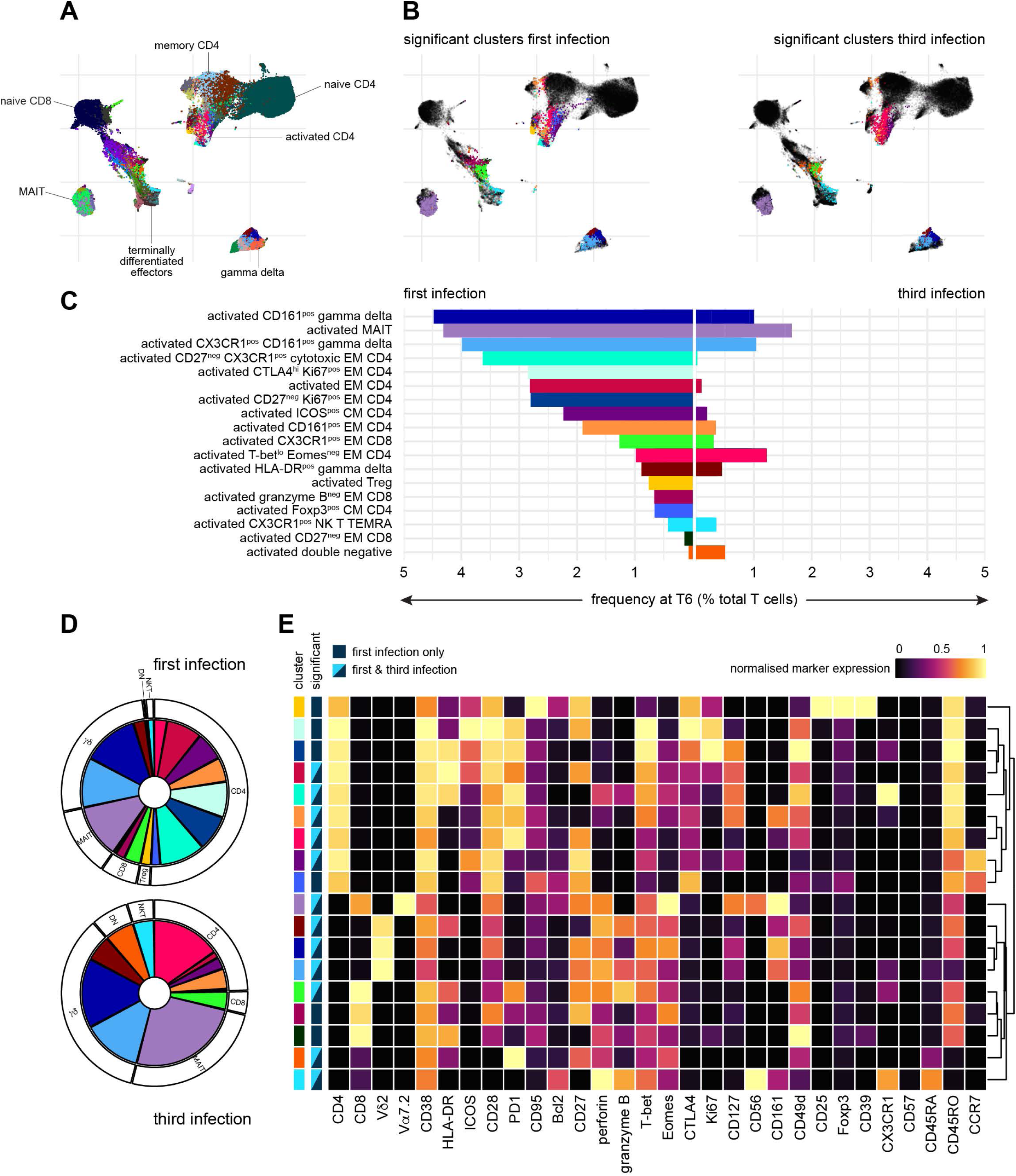
Cytotoxic T cells are silenced for a minimum 8 months. Whole blood was preserved within 30 minutes of blood draw at baseline, diagnosis and T6 during VAC063C as well as 45 days post-challenge (convalescence). Samples were stained with a T cell focussed antibody panel (see Supplementary Table 3) and acquired on a Helios mass cytometer. After exclusion of normalisation beads and doublets we gated on CD45^pos^ CD3^pos^ T cells for downstream steps. (**A**) Data from all volunteers and time-points was concatenated for UMAP analysis and FlowSOM identified 49 discrete clusters of T cells across the dataset (each given a unique colour). The major T cell subsets are labelled according to expression of lineage, memory and activation markers. (**B**) UMAP showing the T cell clusters that are differentially abundant at T6 (versus baseline) in first and third infection (FDR < 0.05 and > 2 fold-change). Clusters that are not significant are shown in black. (**C**) The mean frequency of each T cell cluster that is differentially abundant in first and/or third infection is shown as a proportion of all CD45^pos^ CD3^pos^ T cells at T6. (**D**) Pies show the relative size of each differentially abundant cluster. (**E**) Heatmap showing the normalised median expression values of all markers used for clustering in each of the differentially abundant T cell clusters. The order of features was determined by unsupervised hierarchical clustering. Colour codes to the left of the heatmap indicate cluster identity and show whether clusters were significant in first infection or first and third infection. Note that no cluster was unique to third infection. In (A-E) n = 3 for first infection and n = 6 for third infection.

In first infection eighteen T cell clusters increased in abundance at T6 and all had an activated (CD38^hi^ Bcl2^lo^) phenotype; these were comprised of twelve adaptive and six innate-like clusters that spanned every T cell lineage (Figure 5B-D). In sum, these clusters accounted for approximately 40% of the T cell compartment (Extended Data Figure 6A) and included cytotoxic effectors belonging to the CD8^+^, gamma delta and NK T cell subsets - all were characterised by upregulation of the chemokine receptor CX3CR1 (Figure 5E). CD4^+^ T cells nevertheless dominated this response and we observed enormous heterogeneity with the expansion of nine distinct clusters. There were common traits (such as expression of the memory marker CD45RO) and most had an effector (CCR7^neg^) phenotype but the expression of other activation and differentiation markers (inc. T-bet) was highly variable (Figure 5E). The largest cluster of activated CD4^+^ T cells had an unusual CD27^neg^ CX3CR1^pos^ cytotoxic phenotype and there were other unexpected features of terminal differentiation such as the appearance of perforin-expressing CD161^pos^ effectors. We therefore used limma to complement our analysis of cluster abundance and modelled changes in marker expression through time; this confirmed significant upregulation of both granzyme B and perforin in CD4^+^ T cells six days after drug treatment (Extended Data Figure 6B). Evidently, cytotoxicity is a cardinal feature of the entire T cell response to a first-in-life malaria episode.

In third infection the T cell response was primarily driven by activated innate-like T cells but their abundance was substantially reduced (compared to first infection) and the induction of granzyme B and perforin was suppressed (Figure 5C-D and Extended Data Figure 6B). The number of cytotoxic CD8^+^ T cells was also reduced but the most dramatic change was observed within the CD4^+^ T cell compartment. Here all but one of the nine clusters that expanded in first infection was attenuated and the previously dominant CD27^neg^ CX3CR1^pos^ cytotoxic cluster was completely silenced. These changes were not associated with an increase in regulatory T cells (Foxp3^hi^ CD39^hi^ cells did not expand after re-challenge) or an upregulation of suppressor molecules (Extended Data Figure 6B). In fact, only two clusters increased in size in third compared to first infection - a small subset of double negative T cells and a cluster of T-bet^lo^ Eomes^neg^ effector memory CD4^+^ T cells (Figure 5C and 5E). The majority of activated CD4^+^ T cells belonged to this cluster and in contrast to first infection these cells had already increased in abundance at diagnosis (Supplementary Data File 4); these presumably represent the stem-like memory cells identified by single cell RNA-sequencing. In sum, approximately 10% of the T cell compartment was activated after re-challenge, which more closely aligns with the scale of T cell activation observed in other febrile human infectious diseases.

### Controlling T cell activation protects host tissues

Collectively, these data show that the T cell response to a first-in-life malaria episode is dominated by heterogeneous CD4^+^ T cells that present with unusual features of terminal differentiation and cytotoxicity. To directly test whether T cell activation could be pathogenic we measured biomarkers of collateral tissue damage, a common histological feature of severe malaria. The blood-stage of infection frequently causes liver injury in naive hosts^48, 49, 50^ and auto-aggressive T cells can directly kill primary human hepatocytes^51^ - we therefore measured alanine aminotransferase (ALT) to provide a readout of hepatocellular death *in vivo*. In first infection, two out of three volunteers had abnormal ALT (more than the upper limit of the reference range) when activated T cells were released from inflamed tissues (Extended Data Figure 7A). This was accompanied by increased gamma-glutamyl transferase (GGT) and aspartate aminotransferase (AST) leading to moderate or severe adverse events in both volunteers. In contrast, there was little evidence of any deviation from baseline in liver function tests after re-challenge. To expand our sample size, we performed a meta-analysis using a previously published surrogate dataset; specifically, we examined post-treatment ALT measurements in almost 100 volunteers experiencing a first-in-life infection as part of a human challenge study^49^. Importantly, we only included CHMI trials that were directly comparable to our own re-challenge study - that is they used the same clonal parasite genotype (3D7 or the parental NF54 line); parasites had recently been mosquito transmitted; and similar end-points were applied (treatment at around 10,000 parasites ml^-1^) (Figure 6A). Remarkably, we found that the prevalence of abnormal ALT was reduced from 75% during first infection to 25% upon re-challenge (Figure 6B). And in those rare cases where ALT was increased in second or third infection adverse events were mild (not moderate or severe) even though pathogen load was increased.

**Figure 6.**
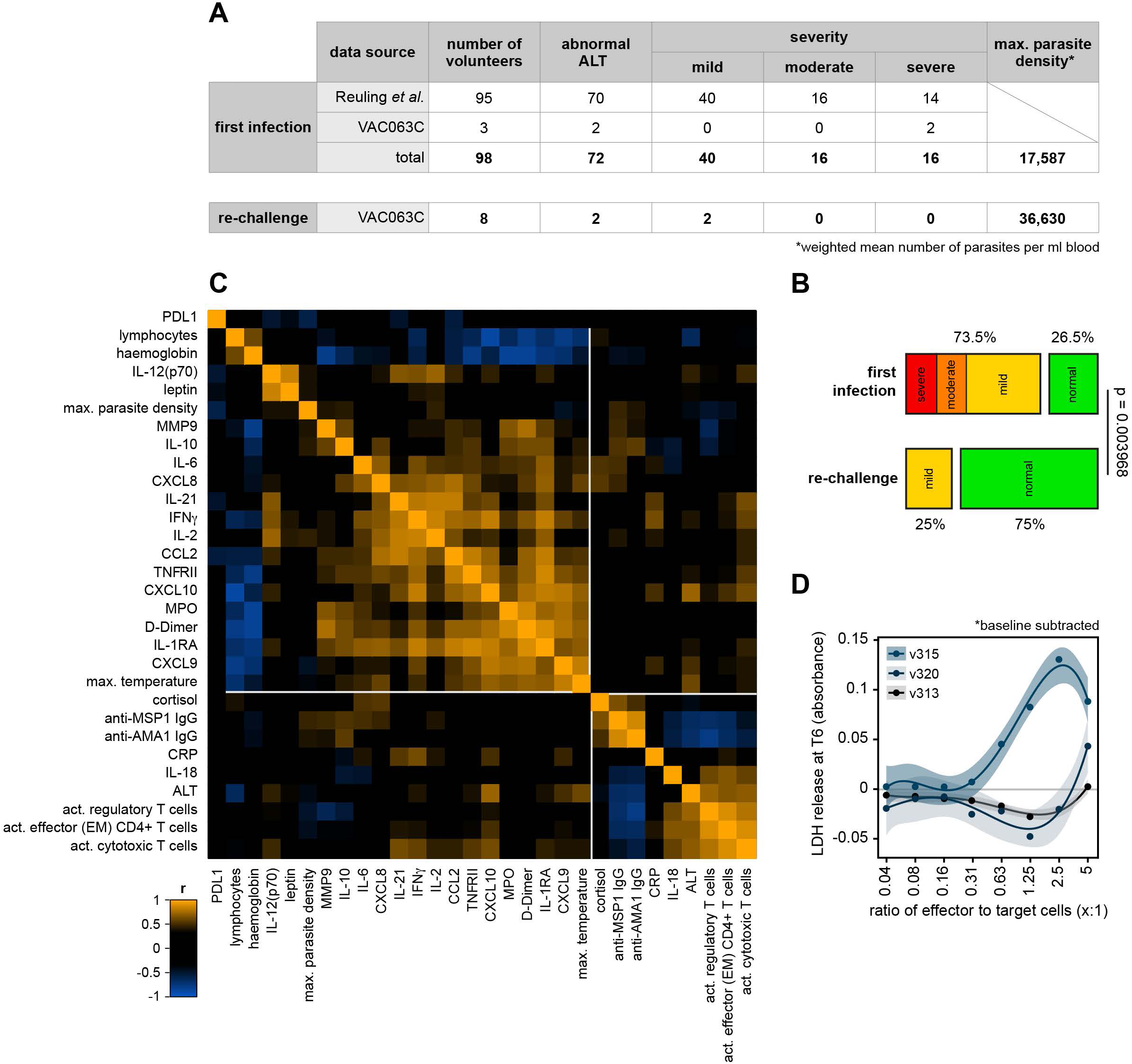
Controlling T cell activation protects host tissues. (**A**) A surrogate dataset from Reuling *et al*.^49^ was used to extract information on the frequency and severity of abnormal alanine aminotransferase (ALT) during a first-in-life infection (up to 6 days post-treatment). All volunteers were infected with *P. falciparum* (3D7 or NF54) as part of a CHMI trial that used equivalent end-points to our own study and in every case abnormal ALT was scored using the same adaptation of the WHO adverse event grading system (see methods). Data from 95 volunteers in Reuling *et al*. (those enrolled in the EHMI-3, LSA-3, EHMI-8B, EHMI-9, ZonMw2, TIP5 and CHMI-trans1 studies) and the 3 first infection volunteers in VAC063C were pooled for analysis. (**B**) Frequency and severity of liver injury; Barnard’s test was used to statistically determine whether an abnormal ALT reading was more prevalent during a first-in-life infection compared to second or third infection (a p value below 0.05 was considered significant). (**C**) Pearson correlation matrix showing the fold-change of differentially abundant plasma analytes, lymphocytes and haemoglobin during VAC063C. Fold-change was calculated either at diagnosis or T6 (relative to baseline) according to when this was largest for each feature. Also included is maximum parasite density, maximum core temperature (up to 48 hours post-treatment) and class-switched antibody titres (28 days after challenge). Finally, circulating ALT is shown at T6 together with the frequency of activated (CD38^hi^) effector (effector memory) CD4^+^ T cells, regulatory T cells and cytotoxic T cells (defined as granzyme B^pos^). All data were log2 transformed and the order of features was determined by unsupervised hierarchical clustering. (**D**) Peripheral blood mononuclear cells (PBMC) were isolated during VAC063C from volunteers undergoing their first infection of life, re-stimulated *in vitro* with PMA/ionomycin and co-cultured with HepG2 cells for 24 hours. Cytotoxicity was measured by the release of lactate dehydrogenase (LDH). Experiments were performed using baseline and T6 samples, and data are shown as baseline subtracted values (i.e. absorbance at T6 minus absorbance at baseline). Curves were fit using a cubic polynomial function (the shaded areas represent 95% confidence intervals). Note that LDH release was specific to HepG2 cells as shown in Extended Data Figure 7B and all assays were run in duplicate. In (A-B) n = 98 for first infection and n = 8 for re-challenge (2 second infection and 6 third infection). In (C) n = 10 (3 first infection, 2 second infection and 5 third infection) and in (D) n = 3 (first infection only).

We then looked more closely at the relationship between T cell activation and liver injury in our cohort. We found a strong positive correlation between ALT and the frequency of activated effector or effector memory CD4^+^ T cells, regulatory T cells and cytotoxic T cells (Figure 6C and Supplementary Table 4). Of note, these all had a strong negative correlation with antibody titres. What’s more, they had no discernible relationship with markers of systemic inflammation or clinical symptoms of malaria, which were instead closely associated with each other but located in a separate clade. Finally, we co-cultured PBMC from first infection with HepG2 cells (an immortalised hepatocyte cell line) and could show enhanced killing at T6 (compared to baseline) *in vitro* (Figure 6D and Extended Data Figure 7B-C). This was particularly prominent for v315 who had by far the largest population of activated CD27^neg^ CX3CR1^pos^ cytotoxic CD4^+^ T cells (Extended Data Figure 6A). Taken together, these data show that the risk of tissue damage and injury can be significantly reduced in the absence of parasite control, which provides the first *in vivo* evidence that long-lived mechanisms of disease tolerance operate in human malaria and can be acquired after a single infection. Moreover, protection does not require the attenuation of systemic inflammation but instead coincides with host control of T cell activation and cytotoxicity.

## Discussion

It has long been recognised that immunity to severe malaria is acquired early in life, offers protection against all manifestations of severe disease and usually precedes clinical immunity (the transition to asymptomatic infection) by more than a decade^2, 3, 4, 5^. We also know that immunity to severe malaria does not require improved parasite control and is thus underpinned by acquired mechanisms of disease tolerance^1^. This is an important distinction to make if we want to understand how to reduce malaria mortality. Host control of inflammation could provide a rapid route to disease tolerance but our data show that malaria parasites trigger a hardwired emergency response across the first three infections of life. This correlates closely with symptomatology in our cohort and is consistent with repeated episodes of fever in children living in endemic settings. In contrast, T cell activation is quickly modified to limit the number and diversity of effector cells, and to avoid cell fates associated with collateral tissue damage and autoinflammatory disease. Our data can therefore begin to explain why large multi-centre genome-wide association studies have repeatedly failed to identify immune loci that associate with severe malaria^52^ - variation in human T cells is almost exclusively driven by non-heritable factors^53^.

If we want to understand how the immune response contributes to severe disease we instead need to ask how malaria triggers such widespread and indiscriminate activation of cytotoxic T cells. These are enriched for markers of terminal differentiation and likely represent short-lived effectors (as evidenced by the downregulation of TCF1 and de-repression of BLIMP1 in CD4^+^ T cells). What remains unclear, however, is whether these are malaria-specific cells or activated via bystander (TCR-independent) mechanisms. If the latter, these could be derived from pre-existing memory cells, which have a far lower threshold for activation than naive cells^54, 55^. This would certainly explain the magnitude of the response, and may also explain the increased susceptibility of adults to severe disease during a first-in-life infection (adults have a larger memory pool than infants and children). Alternatively, malaria may activate autoreactive T cells in the same way that it can activate B cells that produce autoantibodies^56, 57^. Central tolerance (which deletes self-reactive clones in the thymus) is only partially effective and there is enormous degeneracy in TCR reactivity^58^. Systemic infection with a pathogen carrying more than 5000 protein-coding genes may therefore lead to considerable cross-reactivity between parasite and host. In either case, the activation of bystander or cross-reactive T cells would explain our observation that the TCRβ repertoire is essentially unchanged after a first malaria episode.

So how would activation of a large heterogeneous pool of T cells promote severe disease? One possibility is that T cell activation causes extensive activation of the endothelium leading to enhanced parasite cytoadherence and increased pathogen load (an important contributing factor for severe malaria). This has yet to be formally tested but could be assessed *ex vivo* using 3D models of the human microvasculature^59^. This is an important next step because group A/DC8 variants are known to associate with severe disease but do not have an intrinsic growth advantage *in vivo*^14^; a host factor must therefore promote their selection. An alternative (though not mutually exclusive) explanation is that cytotoxic T cells disrupt the endothelial barrier in critical organs and there is emerging evidence for this pathological process in children and adults with cerebral malaria^60, 61^. Furthermore, cytotoxicity may damage parenchymal tissue and our data support a direct role for activated T cells in hepatocyte death. Remarkably, we found that PBMC-mediated killing of HepG2 cells *in vitro* did not require peptide loading and we therefore propose that tissue damage is the result of TCR-independent off-target cytotoxicity. This would explain why we only observe raised transaminases after parasite clearance - activated T cells traffic to the liver during resolution of the immune response to facilitate their clearance^62^. In this scenario, liver injury would be attenuated after re-challenge because the size of the T cell response is diminished. This could be tested *in vivo* using fine needle liver aspirates, which have recently been obtained during CHMI to examine tissue resident memory^63^. In the meantime, we can conclude that explosive cytotoxic T cell activation and tissue damage are the unique features of a first-in-life infection and both are silenced after a single malaria episode to promote tissue health. Importantly, this host adaptation persists for at least 8 months, which would be sufficient to maintain immunity against severe malaria through a prolonged dry season.

It is therefore apparent that we need to start asking how the T cell response to malaria is modified so quickly if we want to obtain a mechanistic understanding of disease tolerance. One possibility is that infection initiates heritable epigenetic programmes that reduce T cell responsiveness. Our data do not provide transcriptional evidence of anergy or exhaustion but this hypothesis needs to be directly tested. An alternative explanation could be suppression by conventional (thymus-derived) regulatory T cells but we found no evidence for their activation in second or third infection - it seems that Tregs are redundant at this point because there is no explosive effector response to keep in check. In much the same way, we found no evidence for the development of IL-10 producing T_R_1 cells, which have been extensively described in endemic settings^64, 65, 66^. At first glance, it could be assumed that this is because T_R_1 cells are generated in the context of chronic stimulation and CHMI is a model of acute infection. However, even in CHMI (where drug treatment is usually initiated within 8 to 12 days of infection) malaria chronically stimulates the immune system. This is because once-infected red cells carry parasite-derived surface antigens and continue to circulate for weeks after drug treatment^67^; follicular dendritic cells present antigens within follicles for months (possibly years)^68^; and hemozoin (an insoluble by-product of infection) persists in lymphoid tissues indefinitely^69^. Indeed, evidence of chronic stimulation can clearly be seen in our study - two clusters of activated (CD38^hi^ Bcl2^lo^) gamma delta T cells are still expanded 45 days after infection (Supplementary Data File 4). It is this persistence of parasite-derived material that we believe is responsible for the long-term protection afforded by a single malaria episode. After all, hemozoin-loaded dendritic cells have a reduced capacity to stimulate T cell proliferation *in vitro*^70^ and a similar mechanism operating within the spleen could provide the simplest explanation for tolerance.

In much the same way, antigen presenting cells can be modified by chronic viral infection and this has been shown to preferentially promote the generation of memory CD8^+^ T cells with stem-like properties^71^. A similar stem-like fate has recently been described for CD4^+^ T cells^72^ and these appear to share remarkable overlap with the TCF1^+^ cluster that is re-activated in our cohort upon re-challenge. TCF1 endows memory CD4^+^ T cells with the capacity for asymmetric division^73^ to promote self-renewal and the differentiation of T_FH_ precursors^37, 74^, which are primed to enter germinal centres or else support B cell responses in the extra follicular pathway. Crucially, asymmetric division is the most efficient way to generate long-lived immunity and the activation of stem-like memory CD4^+^ T cells in third infection (8 months after second infection) coincides with the boosting (MSP-1) and diversification (AMA-1) of parasite-specific class-switched antibodies. A second requirement for long-lived immunity when pathogens persist is reverse TCR evolution, which gradually selects for low affinity T cell clones; this is beneficial because memory cells with a high affinity TCR are driven towards senescence by chronic stimulation^41^. Importantly, effective anti-parasite immunity (and the transition to asymptomatic infection) does not require high affinity antibodies but instead antibody diversification^75, 76, 77^. The polyclonal repertoire of stem-like memory CD4^+^ T cells in third infection may therefore represent the selection of low affinity clones with increased longevity and an enhanced capacity to support a broad repertoire of B cells. As such, we argue that the slow acquisition of anti-parasite immunity in endemic settings (and in our own CHMI study) is not a failure of T cell help.

So do equivalent data from endemic settings support our findings? An important caveat here is that in almost every case samples are collected around the time of drug treatment when activated T cells are still sequestered in inflamed tissues. Nevertheless, we can ascertain that circulating cytotoxic (granzyme B^pos^) CD4^+^ T cells do not expand in children or adults during an uncomplicated episode of febrile disease^78^. In this study, all of these patients would be expected to have acquired immunity to severe malaria because they have lived for at least 2 years in a perennial high transmission setting (183 infective bites per person per year). We can therefore conclude that there is no evidence of cytotoxicity when the risk of severe disease is low. On the other hand, adults with effective anti-parasite immunity have an expanded population of CD161^pos^ central memory CD4^+^ T cells^79^, which may share some similarities with the stem-like memory cells identified here. That’s because stem-like, T_CM_ and T_FH_ cells are all situated on the same branch of the TCF1-BLIMP1 axis (with cytotoxic, T_H1_ and T_R1_ cells on the other side)^37, 38, 74, 80^. And crucially, the frequency of CD161^pos^ T_CM_ CD4^+^ T cells correlates with naturally acquired antibody titres against a broad range of targets (including RH5). These data are therefore consistent with our findings that exposure to malaria quickly silences pathogenic T cells but does not prevent the maintenance of stem-like memory. That this can be achieved through CHMI (a short drug-cured infection) means that interventions that reduce pathogen load but don’t completely eliminate blood-stage parasites could reduce the incidence of clinical malaria in the short-term and have the added benefit or providing long-lived immunity to severe malaria. Moreover, our data indicate that this protection would persist even if these interventions were stopped (drug cover or monoclonal antibodies) or lose efficacy (blood-stage vaccines).

## Methods

### Modelling the risk of severe malaria

To examine the risk of severe disease as a function of prior exposure we used data from a Tanzanian birth cohort^1^. In brief, the authors examined 882 children for *P. falciparum* every 2 - 4 weeks and recorded parasite density and disease severity for each independent infection. Data showing the incidence of severe or complicated malaria (the latter summing severe and moderately severe cases) were extracted from Figure 2C and Supplementary Figure 3, respectively, as shown in the published article. We then used these data to plot the total number of cases of malaria (including mild or uncomplicated episodes); impute missing values by least squares regression (as shown in Extended Data Figure 1); and model risk (as a function of exposure) using the fraction of severe or complicated cases divided by the total number of cases at each order of infection. Binomial regression models were then fit using maximum likelihood methods to test whether the empirical data was best explained by a constant risk (no change), a linear decrease in risk or an exponential decay with increasing exposure. To compare model performance we used the Akaike Information Criterion (AIC) to balance goodness of fit with model complexity, and viewed a reduction in AIC > 2 as indicating a superior model. Maximum likelihood coefficients were estimated with stats4::mle() and stats::glm().

### Clinical trial design - Plasmodium falciparum

All volunteers were healthy malaria-naive adults aged between 18 and 50 years and were enrolled in up to three CHMI studies - VAC063A (November 2017), VAC063B (March 2018) and VAC063C (November 2018). The VAC063A and VAC063B trials evaluated vaccine efficacy of the recombinant blood-stage malaria protein RH5.1 in AS01_B_ adjuvant (GSK) whereas the VAC063C trial investigated the durability of anti-parasite immunity. We collected whole blood samples in each trial from the non-vaccinated (control) volunteers and analysed 8 volunteers in VAC063A (all receiving their first infection), 10 volunteers in VAC063B (8 receiving their second infection and 2 their first infection) and 11 volunteers in VAC063C (6 receiving their third infection, 2 receiving their second infection and 3 their first infection). Our study design and laboratory analysis plan are shown in Supplementary Table 1. All clinical trials received ethical approval from the UK NHS Research Ethics Service - Oxfordshire Research Ethics Committee A for VAC063A and VAC063B (reference 16/SC/0345) and South Central Oxford A for VAC063C (reference 18/SC/0521) - and were registered at ClinicalTrials.gov (NCT02927145 and NCT03906474). VAC063A-C were sponsored by the University of Oxford, carried out in the UK at the Centre for Vaccinology and Tropical Medicine and conducted according to the principles of the current revision of the Declaration of Helsinki 2008 (in full conformity with the ICH Guidelines for Good Clinical Practice). Volunteers signed written consent forms and consent was checked prior to each CHMI. Details of volunteer recruitment, inclusion/exclusion criteria and group allocation can be found in Minassian *et al*. (for VAC063A and VAC063B)^81^ and Salkeld *et al*. (for VAC063C)^82^.

During each CHMI all volunteers were infected with *P. falciparum* (clone 3D7) blood-stage parasites by direct intravenous infusion. The inoculum was thawed and prepared under aseptic conditions and volunteers received between 452 and 857 infected red cells in a total volume of 5 ml saline. Starting one day after challenge volunteers attended clinic every 12 hours for assessment and blood sampling, and parasite density was measured in real-time by qPCR. The parasite multiplication rate was then calculated by fitting linear models to log10 transformed qPCR data, as previously described^83^. In VAC063A thick blood films were evaluated at each time-point by experienced microscopists and diagnosis required volunteers fulfil two out of three criteria: a positive thick blood film (one viable parasite in 200 fields) and/or qPCR data showing at least 500 parasites ml^-1^ blood and/or symptoms consistent with malaria. In VAC063B and C microscopy was dropped to minimise the variation in parasitemia that was observed between volunteers at diagnosis; importantly, this protocol change did not impact volunteer safety and the average parasitemia at diagnosis remained comparable to VAC063A. The new criteria for diagnosis were: asymptomatic with any qPCR result above 10,000 parasites ml^-1^ or symptomatic with a qPCR result above 5000 parasites ml^-1^. In all cases, volunteers were treated with artemether and lumefantrine (Riamet) except where its use was contraindicated and atovaquone and proguanil (Malarone) were given instead. In our analysis, we refer to the blood sample taken immediately before drug treatment (< 30 minutes) as the diagnosis time-point.

Clinical symptoms of malaria (malaise, fatigue, headache, arthralgia, back pain, myalgia, chills, rigor, sweats, nausea, vomiting and diarrhoea) were recorded by volunteers on diary cards or during clinic visits. All symptoms were recorded as adverse events and assigned a severity score: 0 (absent), 1 (transient or mild discomfort), 2 (mild to moderate limitation in activity) or 3 (severe limitation in activity requiring assistance). Pyrexia was scored as absent (≤ 37.5°C), mild (37.6 - 38.2°C), moderate (38.3 - 38.9°C) or severe (≥ 39°C). And full blood counts and blood chemistry (inc. electrolytes, urea, creatinine, bilirubin, alanine aminotransferase, alkaline phosphatase and albumin) were evaluated at the Churchill and John Radcliffe Hospital in Oxford.

### Clinical trial design - Plasmodium vivax

In VAC069A six healthy malaria-naive adults were enrolled to test the infectivity of a new cryopreserved stabilate containing a clonal field isolate of *P. vivax* (PvW1), which had been carefully prepared for use in CHMI^84^. Volunteers were infected with blood-stage parasites by direct intravenous infusion (as above) and the immune response to PvW1 was comprehensively characterised and compared to *P. falciparum* (3D7). In VAC069B three of these volunteers returned for a homologous re-challenge (8-months after VAC069A) and two additional malaria-naive adults received their first challenge with PvW1. In both clinical trials treatment was initiated once two diagnostic criteria were met: a positive thick blood smear, more than 5000 or 10,000 parasites ml^-1^ blood and/or symptoms consistent with malaria. Treatment consisted of artemether and lumefantrine (Riamet) or atovaquone and proguanil (Malarone) if Riamet was contraindicated. Whole blood sampling, processing and downstream analyses were performed analogously to the VAC063 trials (see below). VAC069A and B were sponsored by the University of Oxford, received ethical approval from the UK NHS Research Ethics Service - South Central Hampshire A (reference 18/SC/0577) - and were registered at ClinicalTrials.gov (NCT03797989). The trials were conducted in line with the current version of the Declaration of Helsinki 2008 and conformed with the ICH Guidelines for Good Clinical Practice.

### Processing whole blood for RNA and plasma

Venous blood was drawn into K_2_EDTA-coated vacutainers (BD #367835). To preserve RNA for transcriptional analysis 1 ml whole blood was mixed thoroughly with 2 ml Tempus reagent (ThermoFisher #4342792) and samples were stored at −80°C. No more than 2 hours passed between blood draw and RNA preservation. To obtain platelet-depleted plasma 3 ml whole blood was divided into two 2 ml Eppendorf tubes and centrifuged at 1000 xg for 10 minutes (at 4°C) to pellet cellular components. Working on ice, plasma was then carefully transferred to a new 2 ml tube and centrifuged at 2000 xg for 15 minutes (at 4°C) to pellet platelets. Cell-free platelet-depleted plasma was aliquoted into 1.5 ml Eppendorf tubes, snap frozen on dry ice and stored at −80°C.

### Whole blood RNA-sequencing

RNA was extracted from whole blood using the Tempus spin RNA isolation reagent kit (ThermoFisher #4380204) according to the manufacturer’s instructions. To account for the reduced starting volume and to maintain Tempus stabilizing reagent at the correct final concentration we added just 1 ml PBS to each sample after thawing. Diluted samples were then centrifuged at 3000 xg for 30 minutes (at 4°C) to pellet nucleic acids. Pellets were resuspended in RNA purification resuspension solution and centrifuged on a silica column to remove non-nucleic acid contaminants. After washing, the column was incubated for 2 minutes at 70°C before eluting nucleic acids. 40 μl eluate was then subjected to DNA digestion using the RNA clean and concentrator-5 kit (Zymo Research #R1013). Purified RNA was eluted using 30 μl DNase/RNase-free water, quantified using a Qubit Fluorometer (HS RNA assay kit, ThermoFisher #Q32852) and RNA integrity assessed using an Agilent Bioanalyzer 2100 (RNA 6000 nano kit, Agilent #5067-1511). The average RNA integrity number (RIN) was 9 (98% of samples > 8, lowest RIN 7). Libraries were prepared by Edinburgh Genomics (United Kingdom) using the TruSeq stranded mRNA library prep kit (Illumina #20020595). Stranded library preparation allows transcript expression to be estimated more accurately; in particular it is more effective in quantifying anti-sense gene expression, properly assigning transcripts to putative coding genes and to resolve ambiguity in reads from overlapping genes. Libraries were sequenced using the NovaSeq 6000 Illumina platform yielding 50 bp paired end (PE) reads. These short reads are sufficient to accurately capture gene expression thanks to the well annotated human transcriptome. The average number of reads per sample passing QC across all samples was 8.45 × 10^7^.

### Data analysis of whole blood RNA-sequencing

Quality and content of FASTQ files, which contain raw PE sequencing reads, were assessed using FastQC (http://www.bioinformatics.babraham.ac.uk/projects/fastqc). A single sample (volunteer 1040 baseline second infection) failed quality control and we therefore excluded all RNAseq data from this volunteer’s second infection during analysis. TruSeq primer sequences were removed using Cutadapt v1.9, reads were aligned to the Ensembl release 96 Homo sapiens transcript set with Bowtie2^85^ v2.2.7 (parameters —very-sensitive -p 30 —no-mixed —no-discordant —no-unal) and reads that mapped to globin transcripts were discarded (on average 11.7% of mapped reads, range 1.6 - 38.1%). We opted for bioinformatic globin depletion as it is highly sensitive and reproducible whereas depleting globin during RNA preparation can compromise the amount and quality of RNA recovered^86, 87^. Following these steps, the average alignment rate to the human transcriptome across all samples was 88%. A matrix of normalised counts for each transcript was obtained from sorted, indexed BAM files using Samtools idxstats (http://www.htslib.org/doc/samtools.html). Transcript counts were imported into the R/Bioconductor environment (v3.6) and differential gene expression analyses (pairwise group comparisons) were performed using functions within the DESeq2^22^ package. lfcShrink was applied to the output of each pairwise comparison (type normal) and lists of differentially expressed transcripts were subsequently analysed in R. Non-coding transcripts were removed and multiple transcripts annotated to the same gene were consolidated by keeping the transcript with the highest absolute fold-change. Only protein-coding transcripts with an adjusted p value (adj p) < 0.05 and a fold-change > 1.5 were considered differentially expressed. Volcano/radar/dot plots and heatmaps were generated using the ggplot2^88^ package. The gene lists used for cell cycle analysis were manually compiled from published datasets^89, 90^ and we allocated genes to a single phase based on cell cycle transcriptional network analysis^91, 92^.

### Functional gene enrichment analysis using ClueGO

Lists of differentially expressed genes were imported into ClueGO^93, 94^ v2.5.7. ClueGO identified significantly enriched GO terms (Biological Process and Molecular Function) associated with these genes and placed them into a functionally organised non-redundant gene ontology network based on the following parameters: adj p cutoff = 0.01; correction method used = Bonferroni step down; min. GO level = 5; max. GO level = 11; number of genes = 3; min. percentage = 5; GO fusion = true; sharing group percentage = 40; merge redundant groups with > 40% overlap; kappa score threshold = 0.4; and evidence codes used [All]. Each of the functional groups was assigned a unique colour and a network was then generated using an edge-weighted spring-embedded layout based on kappa score - groups were named by the leading GO term (lowest adj p with min. GO level 5 or 6). Merged networks were constructed by inputting two lists of differentially expressed genes; for each GO term information on what fraction of associated genes were derived from each list was retained. Any GO term containing > 60% associated genes from a single list was considered to be enriched in that group, otherwise GO terms were considered to be shared.

### Multiplexed plasma protein analysis

The concentration of 39 analytes was measured in plasma samples collected at baseline, during infection, diagnosis, 6 days after drug treatment (T6) and 28 or 45 days post-challenge (convalescence). Plasma was thawed on ice and centrifuged at 1000 xg for 1 minute (at 4°C) to remove potential protein aggregates. We customised 4 LEGENDplex panels from BioLegend and performed each assay on filter plates according to the manufacturer’s instructions. Samples and standards were acquired on an LSRFortessa flow cytometer (BD) and FCS files were processed using LEGENDplex software (version 7.1), which automatically interpolates a standard curve using the plate-specific standards and calculates analyte concentrations for each sample. All samples from v1040 were excluded after failing QC and downstream data analysis was performed in R (v3.6). To determine which plasma proteins varied significantly through time we used the lme4 package to fit a separate linear mixed-effects model for each analyte. All available time-points were included and models were fit to log2 transformed data with time-point as a categorical fixed effect and volunteer as a random effect. A Kenward-Roger approximation was used to calculate p values (using the pbkrtest package), which were adjusted for multiple testing using the Benjamini-Hochberg method. Results were visualised with ggplot2 and analytes with at least a 1.5 fold-change from baseline to diagnosis or T6 are shown. To determine which plasma proteins varied between first infection and re-challenge we used lme4 to fit mixed-effects models that included time-point and infection number as categorical fixed effects (with volunteer as a random effect). In this case, linear hypothesis testing was performed using multcomp’s glht function (with Benjamini-Hochberg correction for multiple testing). Only analytes that were significant in both second and third infection (versus first infection) are shown (adj p < 0.05 and fold-change > 1.5).

### Flow-sorting CD4^+^ T cell subsets

We flow-sorted CD4^+^ T cell subsets for bulk RNA-sequencing from 3 ml whole blood collected in K_2_EDTA vacutainers. After red cell lysis (erythrocyte lysis buffer, eBioscience #00-4300-54) leukocytes were washed in PBS supplemented with 2% fetal bovine serum (heat inactivated and 0.22 μm filtered FBS Premium Plus, Gibco #16000044) containing 5 mM EDTA (Life Technologies #AM9260G). We then blocked Fc receptors (human TruStain FcX, BioLegend #422302) and incubated leukocytes for 20 minutes (at 4°C) with the following fluorophore-conjugated antibodies: CD3 (clone OKT3), CD4 (clone OKT4), CD127 (clone A019D5), CD25 (clone M-A251), CCR7 (clone G043H7), CD45RA (clone HI100), CD38 (clone HIT2) and HLA-DR (clone L243) (all from BioLegend). Cells were washed with cold PBS (containing 2% FBS and 5 mM EDTA) and filtered through a 40 μm cell strainer (BD #352340) immediately before sorting. We used a FACSAria III or Fusion cell sorter with a 70 μm nozzle running FACS Diva v8 software (sort setting = purity) to simultaneously sort 10,000 naive (CD127^pos^ CCR7^pos^ CD45RA^pos^), regulatory (CD25^hi^ CD127^neg^) and effector or effector memory (CD127^pos^ CCR7^neg^ CD45RA^neg^) CD4^+^ T cell subsets. Cells were sorted directly into RNase-free sterile 1.5 ml screw cap tubes (ThermoFisher #11529924) containing 1 ml TRIzol Reagent (ThermoFisher #15596026), and incubated for 5 minutes at room temperature. Samples were then stored at −80°C. Note that sort purity was > 95% for every sample - this was assessed in real-time by simultaneously sorting naive CD4^+^ T cells into cold PBS (containing 2% FBS and 5 mM EDTA) and re-acquiring these on the cell sorter.

### Bulk RNA-sequencing of CD4^+^ T cell subsets

RNA was extracted using a modified phenol-chloroform protocol^95^ with 1-Bromo-3-chloropropane (Sigma-Aldrich #B62404) and Isopropanol (Acros Organics #423835000). Total RNA was quantified and integrity assessed using an Agilent Bioanalyzer 2100 (RNA 6000 pico chip, Agilent #5067-1513); all sequenced samples had a RIN value above 8. cDNA was generated from 1 ng total RNA using the SMART-Seq v4 ultra low input RNA kit (Takara Bio #634894) and amplified using 12 cycles of PCR. Amplified cDNA was purified using AMPure XP beads (Beckman Coulter #A63880) and quantified on a Qubit Fluorometer (dsDNA high sensitivity kit, ThermoFisher #Q32851). The quality of the amplified cDNA was then assessed by Bioanalyzer (DNA high sensitivity kit, Agilent #5067-4626). Libraries were constructed from 150 pg cDNA using the Nextera XT DNA library preparation kit (Illumina #FC-131-1024) according to the manufacturer’s instructions. As above, libraries were quantified by Qubit and quality was assessed by Bioanalyser to measure the fragment size distribution. Using this information, samples were combined to create equimolar library pools that were sequenced on a NextSeq 550 Illumina platform to yield 75 bp PE reads; the average number of PE reads per sample passing QC across all samples was 2.74 × 10^7^.

### RNA-sequencing analysis of CD4^+^ T cell subsets

Quality and content of FASTQ files were assessed using FastQC and reads were aligned to the Ensembl release 96 Homo sapiens transcript set with Bowtie2 v2.2.7 (parameters —very-sensitive -p 30 —no-mixed —no-discordant —no-unal). Our median alignment rate was 66.4% after removing primer and adapter traces (SMART-Seq v4 primers, polyG, polyT and polyN) with Cutadapt v1.9. The sorted, indexed BAM files were then used to obtain a matrix of normalised counts for each transcript using Samtools idxstats. Transcript counts were imported into the R/Bioconductor environment (v3.6) and differential gene expression analyses (pairwise group comparisons) were performed using functions within the DESeq2 package. lfcShrink was applied to the output of each pairwise comparison (type normal) and lists of differentially expressed transcripts were filtered by removing all non-coding transcripts and retaining only those with an adj p < 0.05 and fold-change > 1.5. Multiple transcripts annotated to the same gene were consolidated by keeping the transcript with the highest absolute fold-change. Heatmaps and circular stacked bar plots were generated using the ggplot2 package.

### Cryopreservation of peripheral blood mononuclear cells

Peripheral blood mononuclear cells (PBMC) were isolated from 5 ml whole blood, which was diluted with an equal volume of Dulbecco’s phosphate buffered saline (D-PBS) containing 2% fetal bovine serum (FBS) (STEMCELL Technologies #07905) and layered onto 3.5 ml Lymphoprep in a 15 ml SepMate tube (STEMCELL Technologies #07851 and #07905). SepMate tubes were centrifuged at 1200 xg for 10 minutes (at room temperature), the upper plasma-containing layer was discarded and the remaining supernatant above the insert (which contains the PBMC) was poured into a 15 ml tube. Cells were then washed twice in D-PBS containing 2% FBS and cell concentration was determined using a CASY cell counter. Finally, PBMC were resuspended in FBS (heat inactivated and 0.22 μm filtered FBS Premium Plus, Gibco #16000044) containing 10% Dimethyl sulfoxide (DMSO, Sigma-Aldrich #D8418) at a concentration of 5 × 10^6^ cells ml^-1^ and transferred to cryovials. These were placed in a Corning CoolCell freezing container, which was subsequently placed at - 80°C. After 24 hours, PBMC-containing cryovials were transferred to liquid nitrogen where they were stored long-term. To prepare cryopreserved PBMC for use in experiments cryovials were quickly thawed in a 37°C water bath and diluted by adding 10x volume thawing media (RPMI 1640 supplemented with 10% FBS and 20 units ml^-1^ DNase I (Sigma-Aldrich #D4513)) one drop at a time with continuous agitation. Cells were centrifuged at 350 xg for 10 minutes (at room temperature) and washed twice in thawing media. After washing, viable cell counts were calculated using a hemocytometer.

### T cell receptor repertoire sequencing

We analysed the TCRβ repertoire of the six volunteers who underwent third infection during VAC063C, assessing their bulk repertoire prior to challenge (baseline) and 28 days post-challenge (convalescence) during their first (VAC063A) and second (VAC063B) malaria episode. RNA was extracted from thawed PBMC using the Quick-RNA miniprep plus kit (Zymogen) as per the manufacturer’s instructions, including DNase treatment. A modified protocol from Mamedov *et al*.^96^ was then used to synthesise cDNA, which was treated with 1 μl Uracyl DNA glycosylase (5 units μl^-1^). A nested PCR then generated TCRβ V region amplicons with outer primers using indexed forward primers composed of the SMART synthesis oligo sequence fused to a P7 Illumina tag (CAAGCAGAAGACGGCATACGAGATXXXXXXGGCGAAGCAGTGGTATCAACGCAGAGT) and a reverse primer within the TCR C region fused to a P5 Illumina tag (AATGATACGGCGACCACCGAGATCTACACACACSTTKTTCAGGTCCTC). Library QC was performed by Nanodrop and Bioanalyzer, and asymmetric 400 bp + 100 bp sequencing was then performed on an Illumina NovaSeq using custom read primers (ACTCTGCGTTGATACCACTG index with CGAGATCTACACACACSTTKTTCAGGTCCTC for read 1 and GGCGAAGCAGTGGTATCAACGCAGAGT for read 2). The average number of functional TRBV reads was 2.58 × 10^6^ per sample resulting in an average of 13,410 unique CDR3 sequences per sample. Quality control was performed on the raw FASTQ files using FastQC and sequencing data that incorporated UMI was demultiplexed using MiGEC^97^. All sequences were aligned using MiXCR^98^ software utilising IMGT^99^ nomenclature. MiGEC and MiXCR standard error correction thresholds were used (including a minimal nucleotide quality score of 20 within the target gene region) and only in-frame functional CDR3 sequences were included in downstream analyses. Custom pipelines of Python scripts were then used to analyse and plot the MiXCR output and proportional TRBV gene usage was calculated for each sample. Note that for visualisation, TRBV genes that had a zero count at any given time-point were replaced with the minimum gene count prior to log10 transformation.

### Single cell RNA-sequencing of CD4^+^ T cells

Cryopreserved PBMC from volunteers 313, 315 and 320 (first infection) and 1061, 1068 and 6032 (third infection) were prepared 2 days before infection (baseline) and 6 days post-treatment (T6) during VAC063C. To undertake single cell RNA-sequencing these samples were thawed, washed, counted and resuspended at a concentration of 3 × 10^6^ cells ml^-1^ in PBS supplemented with 2% fetal bovine serum (heat inactivated and 0.22 μm filtered FBS Premium Plus, Gibco #16000044) and 5 mM EDTA (Life Technologies #AM9260G). Cells were then stained with surface antibodies exactly as described above (see *Flow-sorting CD4^+^ T cell subsets*) but with the addition of TotalSeq-C oligo-tagged barcoding antibodies that recognise CD298 and β2-microglobulin (a combination of clones LNH-94 and 2M2, available from BioLegend). For each sample we flow-sorted 300,000 CD4^+^ T cells (CD3^pos^ CD4^pos^) into 15 ml conical tubes containing 5 ml RPMI 1640 supplemented with 10% FBS Premium Plus. At the same time we sorted an equal number of CD4^+^ T cells into 5 ml cold PBS (containing 2% FBS and 5 mM EDTA) and re-acquired these on the cell sorter to check purity, which was > 95% for every sample.

After sorting and purity checks, six individually barcoded samples were pooled (one pool for first infection and a separate pool for third infection) and the concentration of each pool was carefully measured and adjusted to 1262 cells µl^-1^. Each pool was then loaded into 2 wells of an A Chip, which itself was loaded onto a 10X Genomics Chromium Controller. Note that we intentionally superloaded the controller in order to capture ∼ 30,000 singlets per pool based on the workflow described in Stoeckius *et al*.^32^ Captured cells were then tagged with 10X cell barcodes in GEMs, cDNA was amplified and libraries were constructed according to the manufacturer’s instructions (Chromium single cell V(D)J reagent kit with feature barcoding technology for cell surface protein - protocol CG000186 revision C). From each GEM three libraries were constructed and separately indexed: i) the cell surface barcode, ii) 5’ gene expression and iii) the T cell receptor ⍺ and β chains (after amplification of the V(D)J regions). Following QC and quantification (using the Agilent Bioanalyzer and TapeStation as well as Qubit) the libraries were pooled at a ratio of 8% cell surface, 84% gene expression and 8% V(D)J and then sequenced on a NovaSeq 6000 Illumina platform to yield at least 66,000 150 bp PE reads per cell. In total we obtained 5931 million PE reads (2848 and 3083 million PE reads for first and third infection, respectively) of which 487 million PE reads (8.2%) were derived from the cell surface library, 4858 million PE reads (81.9%) from the 5’ gene expression library and 586 million PE reads (9.9%) from the V(D)J library.

### Single cell RNA-sequencing analysis

Cell Ranger multi (v6.0.2) was used to align 5’ gene expression and V(D)J sequencing reads to the GRCh38 reference genome. This initial step was performed independently for first and third infection since both pools used the same set of six TotalSeq-C barcodes - C0251 (GTCAACTCTTTAGCG), C0252 (TGATGGCCTATTGGG), C0253 (TTCCGCCTCTCTTTG), C0254 (AGTAAGTTCAGCGTA), C0255 (AAGTATCGTTTCGCA) and C0256 (GGTTGCCAGATGTCA) (all BioLegend). Feature-Barcode matrices were then loaded into R (v4.2.0) using Seurat (v4.3.0) and oligo count matrices were generated from the cell surface library using CITE-seq-Count (v1.4.2) (https://doi.org/10.5281/zenodo.2590196). These datasets were filtered to only include 10X cell barcodes that were detected in both the 5’ gene expression and cell surface libraries, and after normalisation and scaling PCA-initialised tSNE was used to visualise the data (first seven principal components, perplexity = 100). For each pool of samples, inspection of the oligo expression levels revealed six large clusters that corresponded to single positive cells (or singlets). As such, we could use nearest neighbour identification (PCA 1-6) and Louvain clustering to assign each singlet to a sample and exclude cells with multiple oligo tags (doublets). At this stage the data from first and third infection were combined, and a standard Seurat workflow was followed for quality control. Briefly, we excluded cells with more than 2500 feature counts (or less than 200) and cells with more than 5% mitochondrial transcripts. Data were log normalised and the top 2000 most variable features were selected (after variance stabilisation) for computing of the principal components (PC). Harmony^100^ (v0.1.0) was used to integrate data and reduce individual variation, and Seurat’s wrapper for the Louvain algorithm (FindClusters) was used to cluster cells based on the first twelve PC (with default settings at a resolution of 0.8). An initial pass of this analysis identified a persistent and hyperexpanded cluster of TRBV11-3^pos^ CD4^+^ T cells. These cells were unique to v1068 (the only CMV seropositive volunteer) and did not respond to malaria; we therefore excluded this cluster from all downstream steps to avoid confounding signatures of malaria-induced activation.

Next differential cluster abundance was modelled using negative binomial regression with glm.nb() from the MASS package. Time-point was fit as a categorical variable, p values were adjusted (Benjamini-Hochberg method) and an FDR < 0.05 was considered significant. Slingshot^101^ was then used to perform pseudotime analysis on Harmony-derived PC (default settings); the leaf node was set to either cluster 10 or 11 (CD38^hi^ cells) but no trajectory passed through both clusters. Finally, to analyse TCR diversity we used createHTOContigList() to filter and match Cell Ranger’s annotations on the assembled contigs to those extracted from scRepertoire^102^ (v1.8.0). V gene usage (TRAV and TRBV) was analysed using standard immunarch (v0.9.0) (https://github.com/immunomind/immunarch) workflows (excluding ambiguous assignments) and the amino acid sequences of the TCR in each cluster were used to calculate their Gini coefficient (using convenience functions). Data were visualised using ComplexHeatmap^103^ or ggplot2.

### Processing whole blood for mass cytometry

Venous blood was collected in K_2_EDTA-coated vacutainers, stabilised within 30 minutes of blood draw in whole blood stabilisation buffer (Cytodelics #hWBCS002) and stored at −80°C. For antibody staining samples were quickly thawed in a 37°C water bath and then fixed and red cell lysed for 15 minutes using a whole blood preservation kit (Cytodelics #hC002). Next cells were permeabilised with Maxpar barcode permeabilisation buffer (Fluidigm #201057) and each sample was barcoded using Cell-ID 20-plex palladium barcodes (Fluidigm #201060). Samples were then pooled and stained with our T cell focussed surface antibody mix (see Supplementary Table 3) for 30 minutes. After washing, cells were fixed and permeabilised with the Maxpar nuclear antigen staining buffer set (Fluidigm #201063) and incubated with the nuclear antibody mix for 45 minutes. Cells were then washed and fixed for 10 minutes in 1.6% formaldehyde diluted in PBS (ThermoFisher #28906). After a final round of washes, cells were resuspended at a concentration of 3 × 10^6^ cells ml^-1^ in 72.5 nM Cell-ID Intercalator Ir solution (Fluidigm #201192A) and stored overnight at 4°C. Samples were acquired the next day on a freshly tuned Helios mass cytometer (acquisition rate 300-500 events per second) using the WB injector and 10% EQ four element calibration beads (140Ce, 151Eu, 165Ho and 175Lu, Fluidigm #201078).

### Mass cytometry data analysis

The Fluidigm CyTOF software (version 6.7) generated FCS files, which were normalised^104^ and debarcoded^105^ using the R package CATALYST^106^. Samples were compensated using single stained beads^107^. After exclusion of normalisation beads and doublets we gated on CD45^pos^ CD3^pos^ T cells using the Cytobank web portal (https://www.cytobank.org). We then inspected the intensity distribution of each channel and removed those with low variance (CD16, CD69, CXCR5, GATA3, RORγt, TCRγδ and TIM3). The remaining 30 markers were used for UMAP and FlowSOM clustering. UMAP^108^ was used to generate a 2D projection of this high-dimensional dataset. Here phenotypic similarity of cells within and between populations is preserved in the Euclidean distance of the projection. We used its R implementation in the scater^109^ package, which in turn relies on uwot (https://www.github.com/jlmelville/uwot). Features were scaled to unit variance and the 15 nearest neighbours were considered for embedding. UMAP coordinates were then exported for visualisation using ggplot2. FlowSOM^110^ uses self-organising maps (SOM) to efficiently categorise cytometry data into non-overlapping cell populations and was performed using CATALYST (default parameters, target: 100 clusters, 50 metaclusters). After manual inspection we merged two phenotypically similar clusters to avoid overclustering^111^ and ended up with 49 discrete T cell clusters. The R/Bioconductor package ComplexHeatmap was used to visualise T cell phenotypes and the arcsine transformed signal intensity of each marker was independently scaled using a 0-1 transformation across all clusters.

To analyse differential cluster abundance we used the workflow laid out by Nowicka *et al*.^112^ FlowSOM cluster cell counts were modelled linearly with time-point as a dependent categorical variable and volunteer as a fixed effect using the diffcyt^113^ implementation of edgeR^47^. The edgeR functions automatically normalise cluster counts for the total number of cells and improve statistical power by sharing information on cluster count variance between clusters. Pairwise comparisons were performed relative to baseline, and clusters with an FDR < 0.05 and absolute fold-change > 2 were deemed to vary significantly through time. We assessed differential cluster abundance independently for volunteers receiving their first or third infection. We also assessed whether marker expression varied significantly through time and to do this we merged clusters belonging to the same T cell lineage according to their expression of CD4, CD8, CD56, Vδ2 and Vα7.2. Adaptive CD4^+^ and CD8^+^ T cells were then split into naive, effector, effector memory and central memory subsets based on their expression of the markers CD45RA, CD45RO, CD57 and CCR7. All regulatory T cells (CD4^pos^ CD25^hi^ CD127^neg^) were merged into a single cluster. Linear models derived from the limma package, which is optimised for continuous data, were then used to independently assess differential marker expression relative to baseline using pairwise comparisons with moderated t-tests; a shift in median expression of at least 10% and an FDR < 0.05 were required for significance. Results were visualised using ComplexHeatmap with row-wise z-score transformed marker intensities shown for each lineage (or subset) of T cells.

### Meta-analysis of liver injury

A surrogate dataset from Reuling *et al*.^49^ was used to assess the risk of liver injury during a first-in-life infection compared to re-challenge. We extracted data from every CHMI study that used the 3D7 *P. falciparum* clone (or its parental NF54 line), initiated infection via mosquito bite or direct blood challenge and that had a treatment threshold based on thick smear positivity (estimated to be at least 5000 parasites ml^-1^ blood). This produced data for 95 volunteers across 7 CHMI studies. Notably, Reuling *et al*. used longitudinal data to show that liver function test (LFT) abnormalities peaked up to 6 days post-treatment in line with our own T6 time-point. And furthermore, in every CHMI study (including our own) LFT abnormalities were graded using the same adaptation of the WHO adverse event grading system. An LFT reading > 1.0 but < 2.5 times the upper limit of normal was graded as mild; a reading > 2.5 but < 5.0 times the upper limit was graded moderate; and a reading > 5.0 times the upper limit was graded severe. For ALT, the upper limit of normal was 35 or 45 units litre^-1^ for female and male volunteers, respectively. Data from the 95 volunteers in Reuling *et al*. and the 3 volunteers undergoing first infection in our VAC063C study were pooled for analysis and we calculated a weighted peak parasitemia across the cohort by using the mean number of parasites ml^-1^ and the number of volunteers in each of the 8 CHMI studies. To statistically test whether an abnormal ALT reading was more prevalent in the 98 volunteers experiencing their first malaria episode compared to the 8 volunteers undergoing re-challenge (either second or third infection) we used Barnard’s test, which examines the association of two independent categorical variables in a 2 x 2 contingency table. A p value below 0.05 was considered significant.

### Pearson correlation analysis

To assess the relationship between liver injury, T cell activation, systemic inflammation and the clinical symptoms of malaria we constructed a Pearson correlation matrix using our VAC063C dataset. Importantly, this analysis was agnostic of infection number (all volunteers were included). We input the concentration of ALT and the percentage of activated effector (effector memory) CD4^+^ T cells, regulatory T cells and cytotoxic (granzyme B^pos^) T cells (all at T6); the fold-change of lymphocytes, haemoglobin and all significant plasma analytes (those shown in Extended Data Figure 3A), which were calculated at diagnosis or T6 (relative to baseline) according to their largest absolute fold-change; the maximum parasite density and maximum core temperature (at any time-point up to 48 hours post-treatment); and the titres of AMA1/MSP1-specific class-switched antibodies (measured 28 days after challenge). All data were log2 transformed and Pearson correlation was performed in R using the corrplot function. Correlation coefficients were used for unsupervised hierarchical clustering by Euclidean distance.

### In vitro cytotoxicity

The hepatoma cell line HepG2 was kindly provided by Shaden Melhem (University of Edinburgh). HepG2 cells were maintained at 37°C and 5% CO_2_ in RPMI 1640 supplemented with 10% heat-inactivated fetal bovine serum (FBS Premium Plus, Gibco #16000044), 1 x GlutaMAX (Gibco #35050061), 100 units ml^-1^ penicillin and 100 µg ml^-1^ streptomycin (Gibco #15140122). Expansion was performed in tissue culture treated culture flasks (Corning #430641U) and cells were detached using TrypLE Express Enzyme (Gibco #12604013). For cytotoxicity assays, HepG2 cells were seeded at a density of 5.5 × 10^5^ cells per well in 96 well tissue culture treated plates pre-coated with Poly-D-Lysine (Gibco #A3890401) and incubated for 24 hours to allow formation of a confluent monolayer.

The next day PBMC were thawed, washed, counted and resuspended at a concentration of 3 × 10^6^ cells ml^-1^ in RPMI 1640 supplemented with 10% fetal bovine serum (FBS Premium Plus), 2 mM L-Glutamine (#25030081), 1 mM sodium pyruvate (#11360070), 10 mM HEPES (#15630080), 500 µM β-mercaptoethanol (#31350010) and 1 x MEM non-essential amino acids (#M7145) (all Gibco except amino acids from Sigma-Aldrich). Cells were transferred to ultra low attachment plates (Corning #3473) and stimulated with cell activation cocktail (BioLegend #423302) for 3 hours at 37°C and 5% CO_2_ - the working concentration of PMA and ionomycin used was 40.5 nM and 669.3 nM, respectively. PBMC were then harvested, washed twice and added onto HepG2 monolayers at increasing ratios of effector (PBMC) to target cells (HepG2). After 24 hours of co-culture the release of lactate dehydrogenase (LDH) from damaged HepG2 cells was measured in the culture supernatant using the CyQUANT LDH cytotoxicity assay according to the manufacturer’s instructions (Invitrogen #C20301). Background LDH release was quantified in wells containing only HepG2 cells. Absorbance was read on a FLUOstar Omega plate-reader.

## Acknowledgements

We thank Paul Sopp for T cell sorting during the VAC063 trial and Neil Ashley for support with single cell RNA-sequencing. Flow-sorting, 10X Genomics and CyTOF data were generated within the flow cytometry, single cell and mass cytometry facilities at the Weatherall Institute of Molecular Medicine (University of Oxford), which are supported by MRC Human Immunology Unit core funding (MC_UU_00008) and the Oxford Single Cell Biology Consortium (OSCBC). CD4^+^ T cell subset RNAseq libraries were prepared and sequenced by the Edinburgh Clinical Research Facility at the University of Edinburgh, which receives financial support from NHS Research Scotland (NRS). Whole blood RNAseq libraries were prepared and sequenced by Edinburgh Genomics, which is supported through core grants from NERC (R8/H10/56), MRC UK (MR/K001744/1) and BBSRC (BB/J004243/1). We would like to extend our thanks to the VAC063 and VAC069 clinical and laboratory teams for assistance, and to all of the volunteers who participated in this study.

## Data availability

All bulk RNAseq data (whole blood and sorted T cell subsets) have been deposited in NCBI’s Gene Expression Omnibus and are accessible through GEO SuperSeries accession number GSE172481. Single cell RNA-sequencing data have also been deposited in NCBI’s Gene Expression Omnibus and are available through accession number GSE275092. Bulk TCRβ sequencing data have been deposited in the European Nucleotide Archive and are available through accession number PRJEB71976. And CyTOF (mass cytometry) data have been deposited at flowrepository.org - these can be accessed through experiment numbers FR-FCM-Z47Z (VAC063C); FR-FCM-Z3HA (VAC069A); and FR-FCM-Z465 (VAC069B).

---

**Extended Data Figure 1.**
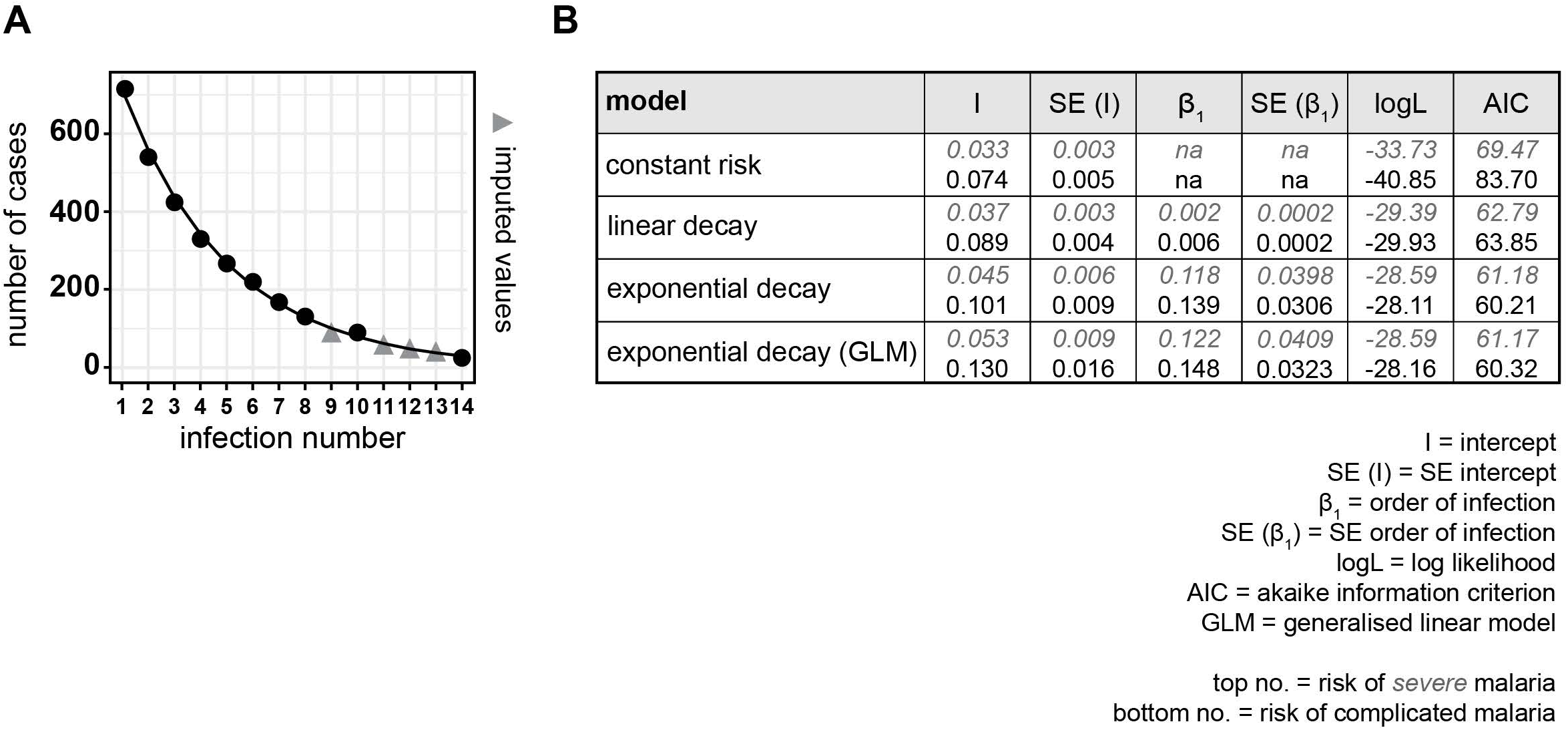
The risk of severe malaria decreases exponentially with exposure. Data were extracted from Gonçalves *et al*.^1^ to examine the frequency of severe or complicated malaria during the first 14 infections of life in infants living in a hyperendemic setting. (**A**) We first plotted the total number of cases of malaria (including mild or uncomplicated episodes) and used least squares regression to impute missing values (for infection number 9, 11, 12 and 13). Imputed values are shown as a grey triangle whereas filled circles indicate data as reported by Gonçalves *et al*. The total number of children experiencing at least 1 episode of malaria was n = 715. (**B**) We then plotted the incidence of severe or complicated malaria at each order of infection (as shown in Figure 1A-B) and performed maximum likelihood estimation to select the best model fit for these data; log likelihood (logL) and AIC both show that an exponential decay in risk provides the best fit.

**Extended Data Figure 2.**
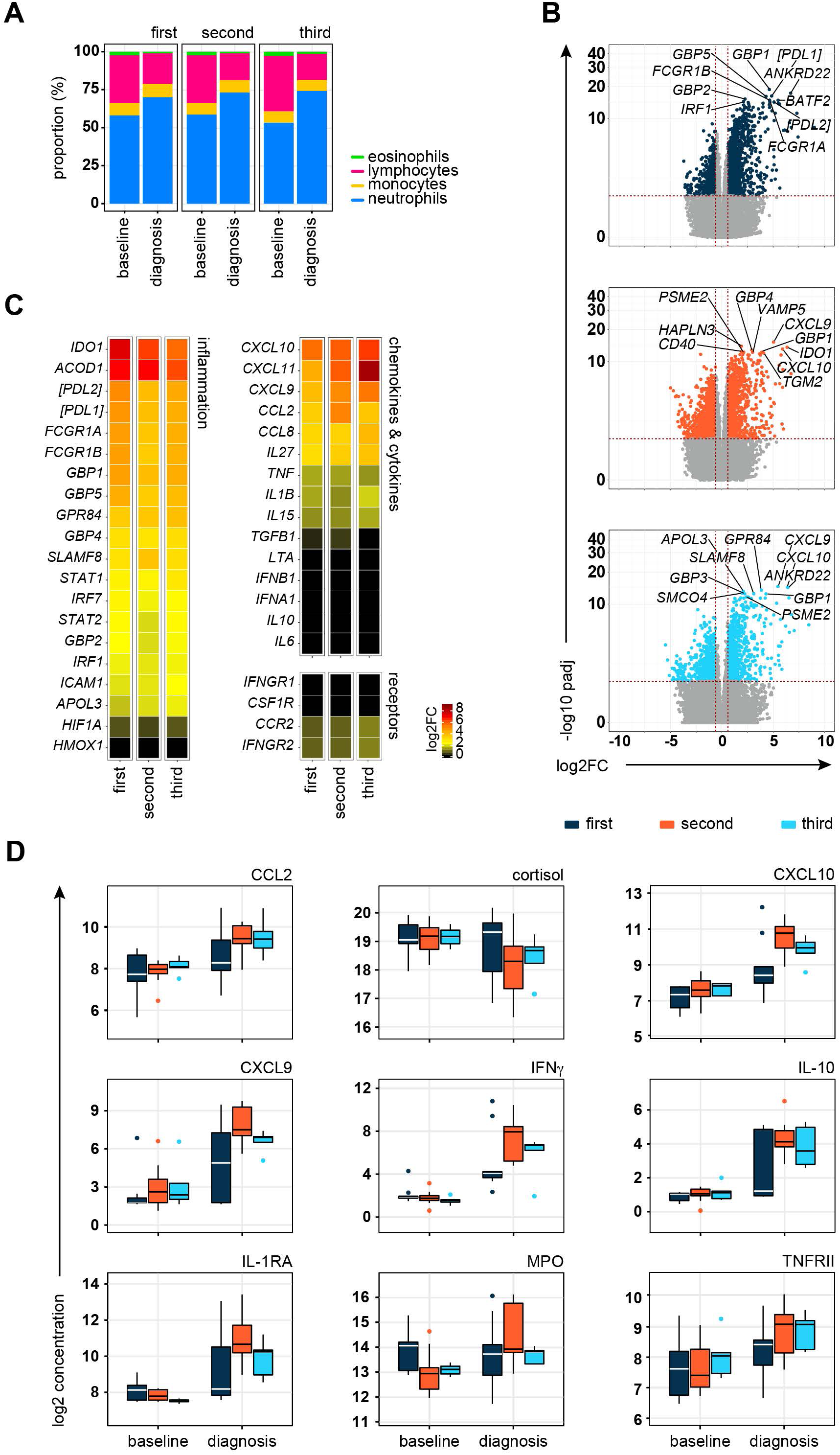
Systemic inflammation is not attenuated upon re-challenge. (**A**) Proportion of eosinophils, lymphocytes, monocytes and neutrophils in whole blood at baseline and diagnosis. The mean frequency is shown for each time-point and infection. Note that the loss of lymphocytes at diagnosis is comparable in all three infections. (**B**) RNA-sequencing was used to identify differentially expressed genes in whole blood at diagnosis (versus baseline) (adj p < 0.05 and > 1.5 fold-change). Volcano plots show all differentially expressed genes (coloured dots) and the dashed lines represent the significance/fold-change cut-offs (genes that are not significant are shown in grey). The top 10 differentially expressed genes (lowest adj p) in each infection are labelled (first, second and third infection were analysed independently). (**C**) The log2 fold-change of signature genes associated with interferon signaling and type I inflammation are shown at diagnosis (versus baseline) in first, second and third infection. Square brackets indicate that common gene names have been used. (**D**) Linear regression was used to identify plasma analytes that vary significantly between infections at diagnosis. Multiple test correction was performed using the Benjamini-Hochberg method and analytes that were significant in both second and third infection (versus first infection) are displayed (adj p < 0.05). Box (median and IQR) and whisker (1.5x upper or lower IQR) plots are shown with outliers as dots. In (A) n = 10 (first and second infection) and n = 6 (third infection). In (B-C) n = 10 (first infection), n = 9 (second infection) and n = 6 (third infection). v1040 was excluded from RNA-sequencing analysis in second infection because their baseline sample failed QC. In (D) n = 9 (first and second infection) and n = 5 (third infection). v1040 was excluded from plasma analysis because all samples failed QC.

**Extended Data Figure 3.**
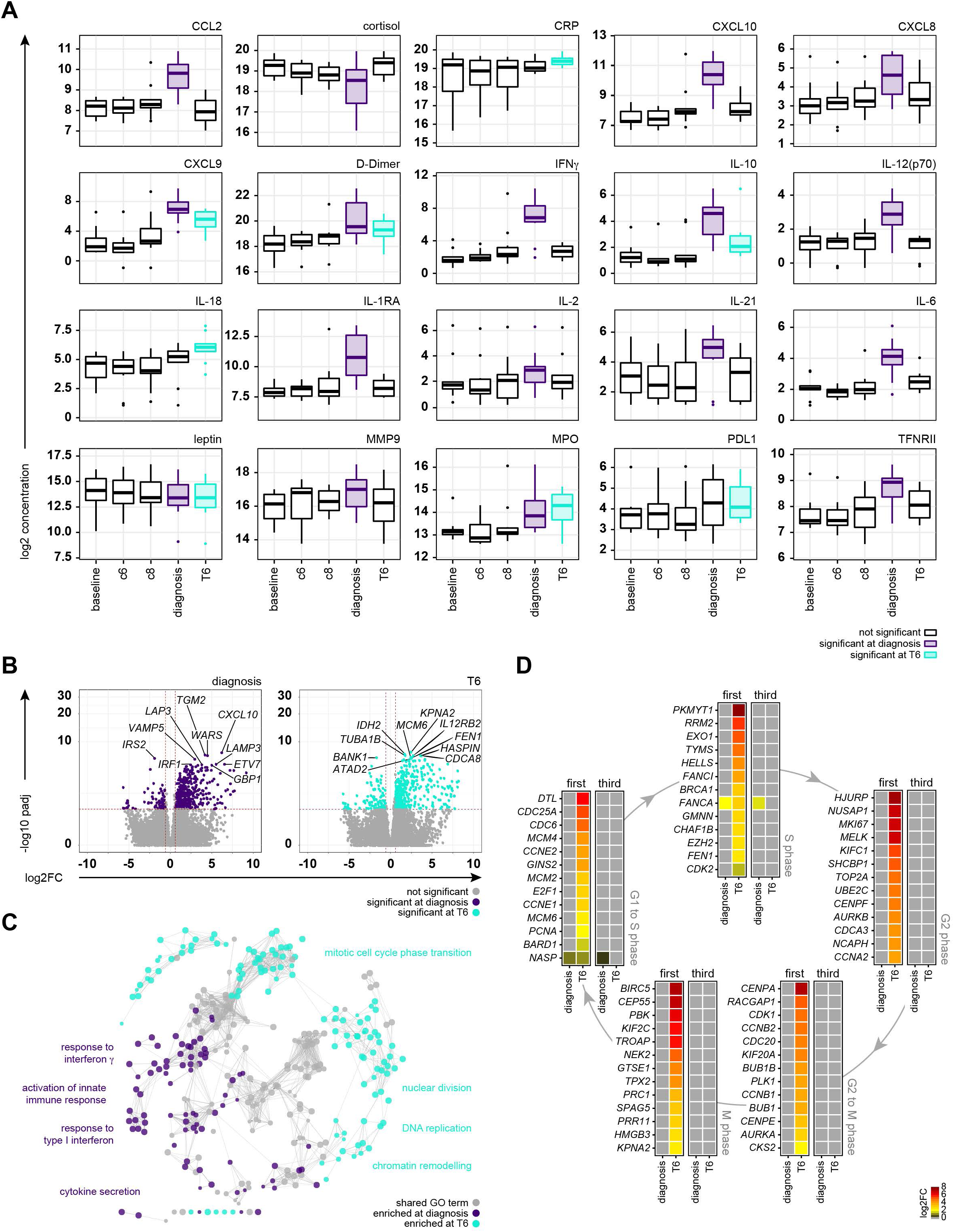
The acute phase response is followed by proliferation in first infection. (**A**) Mixed-effects models and linear regression were used to identify plasma analytes that vary significantly at diagnosis and/or T6 across the entire VAC063C dataset (all volunteers regardless of infection number). Kenward Roger approximation was used to calculate p values and multiple test correction was performed using the Benjamini-Hochberg method; significance (adj p < 0.05) is indicated by coloured box plots (purple at diagnosis and turquoise at T6). Box (median and IQR) and whisker (1.5x upper or lower IQR) plots are shown with outliers as dots. (**B**) RNA-sequencing was used to identify differentially expressed genes in whole blood at diagnosis and T6 during first infection (versus baseline) (adj p < 0.05 and > 1.5 fold-change). Volcano plots show all differentially expressed genes (coloured dots) and the dashed lines represent the significance/fold-change cut-offs (genes that are not significant are shown in grey). The top 10 differentially expressed genes (lowest adj p) at each time-point are labelled. (**C**) Differentially expressed genes at T6 and diagnosis were combined for GO analysis of first infection and ClueGO was used to construct a merged functional gene ontology network. Each node represents a GO term and nodes are coloured according to whether their associated genes were majoritively (> 60%) derived from the diagnosis or T6 time-point. GO terms that were shared between time-points are coloured grey (see methods). Four leading GO terms (each from a unique functional group) are labelled for each time-point. (**D**) Heatmaps show differentially expressed genes at diagnosis and T6 (versus baseline) during first and third infection (adj p < 0.05 and > 1.5 fold-change). The log2 fold-change of key genes associated with each phase of the cell cycle is shown. Non-significant genes are displayed with a log2 fold-change of zero. In (A) n = 10 (3 first infection, 2 second infection and 5 third infection). v1040 was excluded from plasma analysis because all samples failed QC. In (B - C) n = 3 (first infection only) and in (D) n = 3 (first infection) and n = 6 (third infection).

**Extended Data Figure 4.**
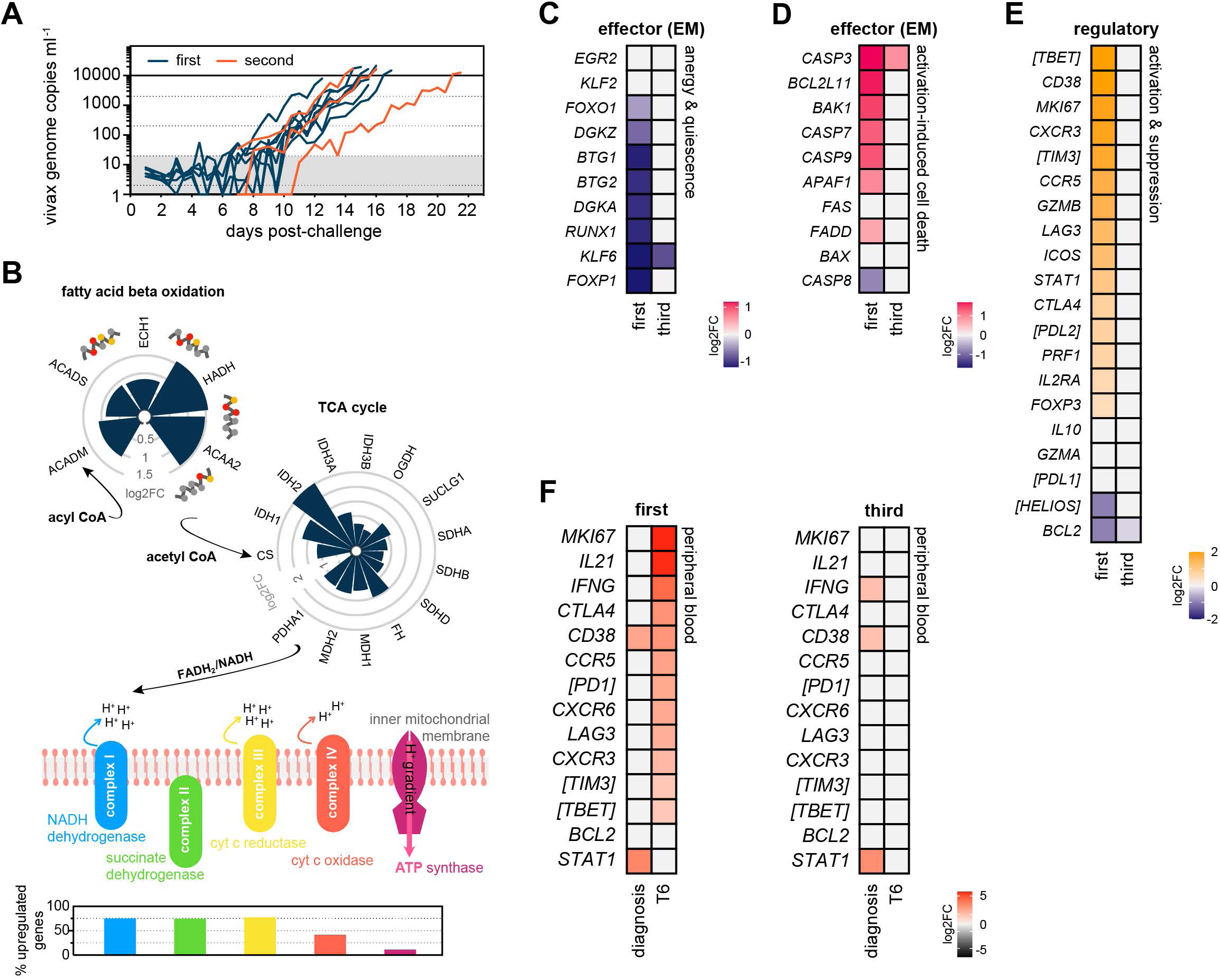
Re-challenge does not induce CD4^+^ T cell anergy, exhaustion or clonal deletion. (**A**) Healthy malaria-naive adults were infected up to two times with *P. vivax* (clone PvW1) by direct blood challenge during the VAC069 study. Parasite growth curves are shown for first and second infection; each line represents a volunteer and lines are colour-coded by infection number. Parasite density was measured in peripheral blood by qPCR every 12 - 24 hours. The grey box represents the lower limit of quantification (20 parasites ml^-1^) and the treatment threshold of 10,000 parasites ml^-1^ is denoted by the black line. (**B**) RNA-sequencing was used to analyse transcriptional regulation of fatty acid β-oxidation, the tricarboxylic acid (TCA) cycle and oxidative phosphorylation (oxphos) in flow-sorted effector (effector memory) CD4^+^ T cells during first infection with *P. falciparum* (VAC063C). The circular bar charts show the log2 fold-change of each major enzyme involved in fatty acid β-oxidation and the TCA cycle clockwise in reaction order 6 days after parasite clearance (T6 versus baseline). The vertical bar chart shows the proportion of oxphos enzymatic subunits that are transcriptionally upregulated at T6 - all subunits required to form complex I to IV in the electron transport chain and ATP synthase are shown. The key molecules that connect these metabolic pathways are labelled. (**C** - **D**) RNA-sequencing was used to identify differentially expressed genes in flow-sorted effector (effector memory) CD4^+^ T cells in first and third infection during VAC063C (T6 versus baseline) (adj p < 0.05 and > 1.5 fold-change). Heatmaps show the log2 fold-change of (C) hallmark genes indicating T cell anergy or quiescence and (D) markers of activation-induced cell death. (**E**) Flow-sorted regulatory T cells (CD4^pos^ CD25^hi^ CD127^neg^) were also analysed by RNA-sequencing to identify differentially expressed genes relating to their activation and suppressor function (T6 versus baseline) in first and third infection (adj p < 0.05 and > 1.5 fold-change). (**F**) RNA-sequencing was used to identify differentially expressed genes in whole blood at diagnosis and T6 (relative to baseline) in first and third infection during VAC063C (adj p < 0.05 and > 1.5 fold-change). Signature genes associated with T cell activation, T_H_1 differentiation and cytokine production are shown. In (A) n = 8 for first infection and n = 3 for second infection. In (B - E) n = 2 or 3 for first infection (T6 and baseline, respectively) and n = 6 for third infection (v313 was excluded at T6 because this sample failed QC). In (F) n = 3 for first infection and n = 6 for third infection. In (C-F) non-significant genes are displayed with a log2 fold-change of zero.

**Extended Data Figure 5.**
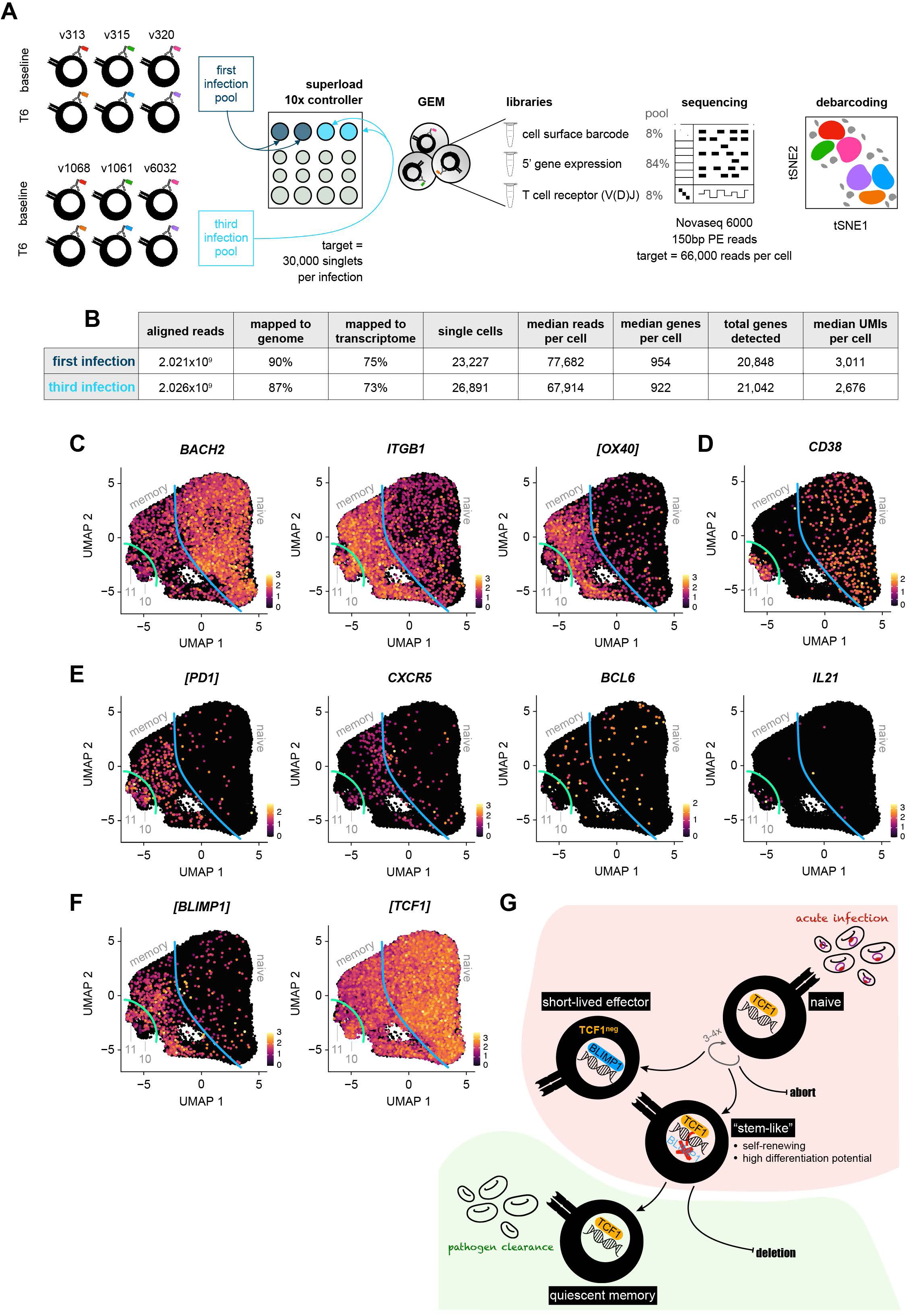
Activated CD4^+^ T cells bifurcate along the TCF1/BLIMP1 axis. (**A**) VAC063C single cell RNA-sequencing workflow: each sample of flow-sorted CD4^+^ T cells was barcoded using TotalSeq-C oligo-tagged antibodies; samples from all volunteers and time-points were pooled (separately for first and third infection); and pooled samples were superloaded onto a 10X Chromium Controller (we aimed to capture 30,000 singlets per pool). Gel beads in emulsion (GEMs) encapsulating a single cell (or doublets) were then generated and from each GEM three libraries were produced: i) the cell surface barcode, ii) 5’ gene expression and iii) T cell receptor (after amplification of the V(D)J regions). Libraries were pooled at the specified ratios and sequenced. Finally, we used PCA-based clustering to debarcode all samples and remove doublets (see methods). (**B**) Cell Ranger was used to align 5’ gene expression and V(D)J sequencing reads (independently for first and third infection). Shown is the output of Cell Ranger after removing doublets and performing QC. (**C** - **F**) Data from all volunteers and time-points was concatenated for UMAP analysis. The expression intensity of markers for memory (C), activation (D) and T_FH_ differentiation (E) are shown across the UMAP. The blue line represents the split between naive and memory cells whereas the green line represents the split between memory and activated cells. In (F) the expression intensity of the master transcription factors associated with terminal differentiation (BLIMP1) versus the maintenance of stem-like properties (TCF1) are shown. In all cases, each UMAP is equivalent to those shown in Figure 4 (for cross-reference) and square brackets indicate that common gene names have been used. (**G**) Proposed model of T cell activation during a first-in-life malaria episode. The maintenance of stem-like T cells is essential for long-lived memory; this requires sustained expression of TCF1 to repress BLIMP1 and prevent the terminal differentiation of short-lived effector cells. In (A - F) n = 3 for first and third infection.

**Extended Data Figure 6.**
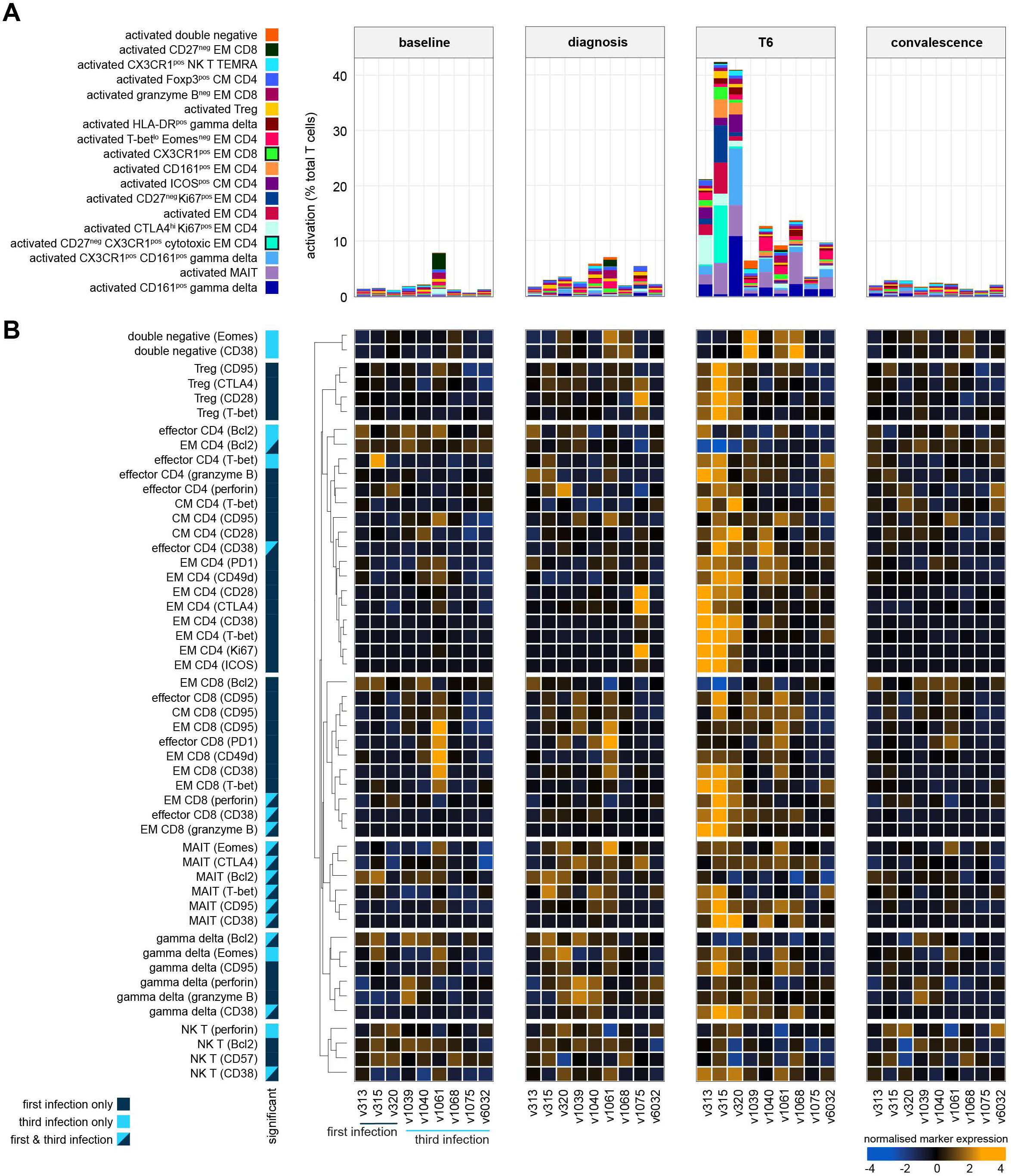
Cytotoxic T cells are silenced after a single malaria episode. (**A**) Stacked bar chart showing the frequency of activated (CD38^hi^ Bcl2^lo^) T cells at baseline, diagnosis and T6 as well as 45 days post-challenge (convalescence) during VAC063C. Each bar represents one volunteer and individual T cell clusters are colour-coded to match Figure 5 (note that only differentially abundant clusters are included). The major CD4^+^ and CD8^+^ T cell clusters with cytotoxic features are highlighted with a black border in the key to the left of the plot. (**B**) Differential marker expression through time in each major T cell subset. First, T cell clusters belonging to the same lineage were merged and then CD4^+^ and CD8^+^ T cells were split into naive, effector, effector memory (EM) and central memory (CM) subsets. Next, linear models were used to independently assess differential marker expression in each subset at each time-point (relative to baseline); a shift in median expression of at least 10% and an FDR < 0.05 were required for significance. Shown are all subset/marker pairs that were called as significant at T6 and data are presented as row-wise z-score marker intensities. Colour codes to the left of the heatmap indicate whether markers were differentially expressed during first infection, third infection or both infections.

**Extended Data Figure 7.**
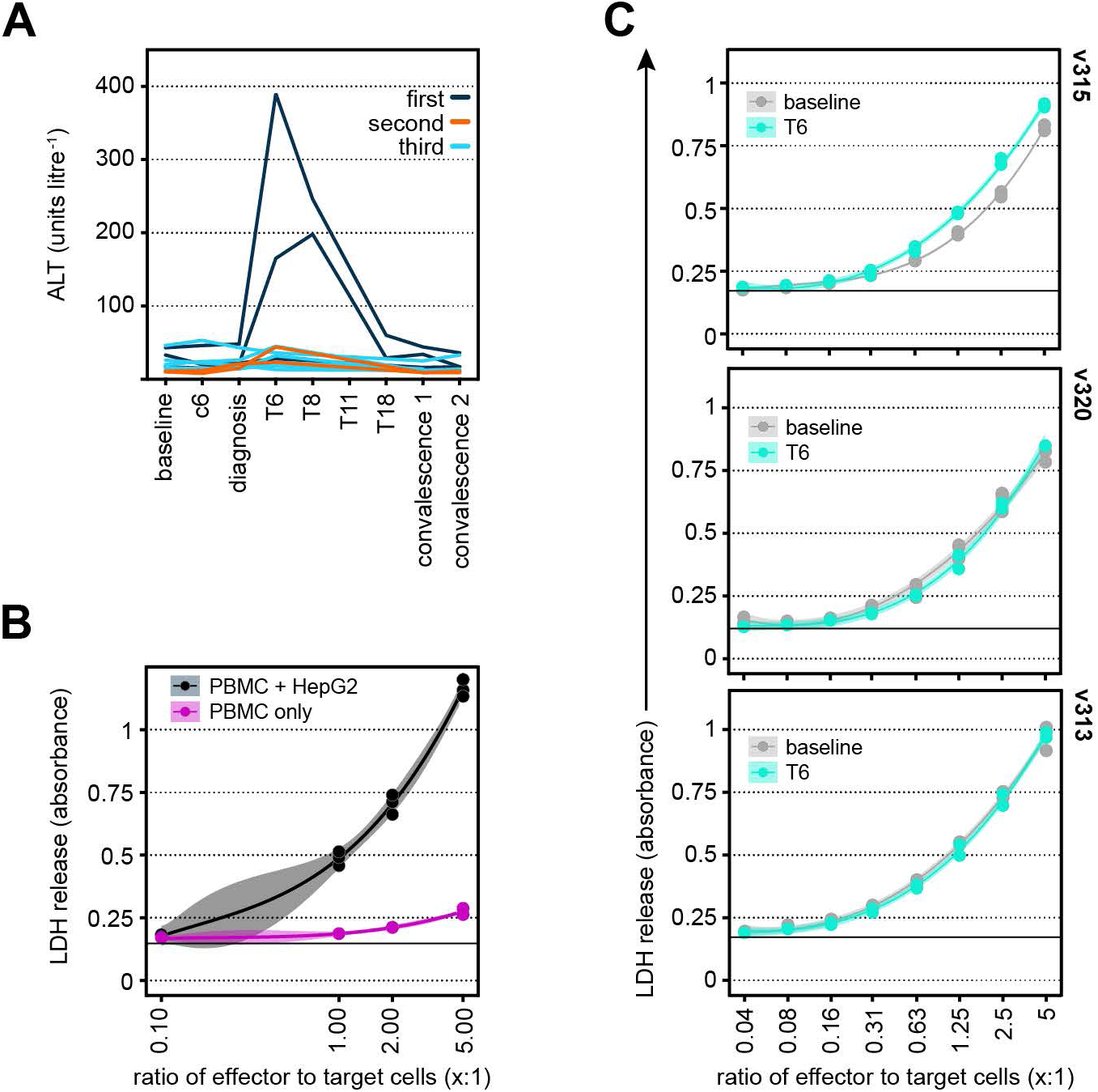
Raised serum transaminases are unique to first infection. (**A**) Blood chemistry measured the concentration of alanine aminotransferase (ALT) through infection and convalescence in the VAC063C study (baseline, 6 days post-challenge (c6), diagnosis, 6 - 18 days after drug treatment (T6 - T18) and 45 or 90 days post-challenge (convalescence 1 and 2, respectively)). Each line represents one volunteer and lines are colour-coded by infection number. (**B** - **C**) Peripheral blood mononuclear cells (PBMC) were isolated during VAC063C from volunteers undergoing their first infection of life, re-stimulated *in vitro* with PMA/ionomycin and co-cultured with HepG2 cells for 24 hours. Cytotoxicity was measured by the release of lactate dehydrogenase (LDH). (B) PBMC were co-cultured with or without HepG2 cells to determine their relative contribution to LDH release. (C) PBMC isolated at baseline (grey) or T6 (turquoise) were co-cultured with HepG2 cells at different effector to target cell ratios. Each plot represents the raw data from one volunteer, the black line shows the mean absorbance in control wells (HepG2 cells only) and volunteers are ordered according to their frequency of activated cytotoxic CD4^+^ T cells at T6 (highest to lowest (top to bottom) - see Extended Data Figure 6A). In (A) n = 11 (3 first infection, 2 second infection and 6 third infection). In (B) a representative experiment is shown and in (C) n = 3 (first infection only). Note that all cytotoxicity assays were performed in duplicate or triplicate and curves were fit using a cubic polynomial function (the shaded areas represent 95% confidence intervals).

**Supplementary Data File 1.** Gating strategy for sorting CD4^+^ T cell subsets during VAC063C. CD4^+^ T cells were sorted *ex vivo* (within 2 hours of blood draw) into TRIzol for downstream RNA-sequencing - cells with a naive, effector (effector memory) or regulatory phenotype were sorted as shown at baseline and T6. Note that we did not use CD38 for sorting but subsequently used this marker to assess the level of activation within each subset at both time-points.

**Supplementary Data File 2.** T cell receptor V gene usage in CD4^+^ T cell clusters. Droplet-based single cell RNA-sequencing was carried out during VAC063C on flow-sorted CD4^+^ T cells obtained at baseline and T6 from volunteers undergoing their first or third infection of life. Data from all volunteers and time-points was concatenated to examine V gene usage (in TCR⍺ and TCRβ chains) in each cluster. Note that clusters are colour-coded exactly as shown in Figure 4 and it is clusters 10 (self-renewal) and 11 (terminal differentiation) that are activated (CD38^hi^) by malaria (n = 3 in first and third infection).

**Supplementary Data File 3.** T cell cluster phenotypes by mass cytometry. Heatmap showing the normalised median expression values of all markers used for clustering in each of the 49 T cell clusters. Names were assigned manually using activation, lineage and memory markers to broadly categorise each T cell cluster; when more than one cluster was placed into the same category (e.g. activated EM CD4) clusters were given an accessory label to highlight their unique phenotype or property (e.g. T-bet^lo^ Eomes^neg^). The order of features was determined by unsupervised hierarchical clustering.

**Supplementary Data File 4.** T cell cluster frequencies by mass cytometry. Each cluster is shown as a proportion of all CD45^pos^ CD3^pos^ T cells at each time-point. Clusters are shown in the same order as Supplementary Data File 3 (left to right and top to bottom). Box (median and IQR) and whisker (1.5x upper or lower IQR) plots are shown (with outliers as dots) and significance (FDR < 0.05 and > 2 fold-change) is indicated by colour (dark blue for first infection and bright blue for third). In all plots n = 3 for first infection and n = 6 for third infection.

**Supplementary Table 1.** Demographics of volunteers infected and re-challenged with *Plasmodium falciparium* (3D7) during VAC063A-C; includes genetic and non-genetic variables known to influence human immune variation *in vitro*. Also shown is the parasite multiplication rate (detailed methodology for the modelling of qPCR data can be found in Minassian *et al*.^81^) and maximum circulating parasite density. There were no statistically significant differences between first, second and third infection (Kruskal-Wallis test, p = 0.3240 for parasite multiplication rate and p = 0.5975 for maximum parasite density).

**Supplementary Table 2.** Signature genes associated with terminal differentiation and self-renewal in CD4^+^ T cells. Tab one lists the signature genes for each of the 13 clusters of CD4^+^ T cells identified by single cell RNA-sequencing during VAC063C. Note that clusters are colour-coded exactly as shown in Figure 4 and for each cluster genes are ordered by adj p value. Tab two lists the differentially expressed genes identified by direct pairwise comparison between activated (CD38^hi^) clusters 10 and 11. Genes that are more highly expressed in cluster 10 have a positive log2 fold-change. Note that differential gene expression analysis was performed across the entire dataset (i.e. data from all volunteers and time-points was concatenated) (n = 3 in first and third infection).

**Supplementary Table 3.** Mass cytometry antibody panel for T cell fate and function in VAC063C; includes information on antibody clone, source and heavy metal conjugate.

**Supplementary Table 4.** Coefficient and p values underlying the Pearson correlation matrix shown in Figure 6C.

